# A comprehensive systematic review and meta-analysis of the global data involving 61,532 cancer patients with SARS-CoV-2 infection

**DOI:** 10.1101/2021.12.18.21267261

**Authors:** Emma Khoury, Sarah Nevitt, William Rohde Madsen, Lance Turtle, Gerry Davies, Carlo Palmieri

**Affiliations:** University of Liverpool, Institute of Translational Medicine, Department of Molecular and Clinical Cancer Medicine, UK; University of Liverpool, School of Medicine, Liverpool, UK; Department of Health Data Science, Institute of Population Health, University of Liverpool, UK; Department of Political Science & School of Public Policy, University College London; University of Copenhagen, Department of Political Science; Tropical and Infectious Disease Unit, Liverpool University Hospitals NHS Foundation Trust, member of Liverpool Health Partners; University of Liverpool Department of Clinical Infection Microbiology and Immunology, Department of Clinical Infection, Liverpool; University of Liverpool Institute of Infection and Global Health, Veterinary and Ecological Sciences, Liverpool; The Clatterbridge Cancer Centre NHS Foundation Trust, Liverpool, UK

**Keywords:** SARS-CoV-2, COVID-19, Cancer, Systematic Review

## Abstract

**Background:** SARS-CoV-2 have been shown to be associated with more severe disease and death in cancer patient. A systematic review and meta-analysis was conducted to determine the risk by age, tumour type and treatment of infection with SARS-CoV-2 in cancer patients.

**Methods:** Systematic review by searching PubMed, Web of Science, and Scopus for articles published in English up to June 14, 2021 of SARS-CoV-2 infection in >10 patients with malignant disease. Outcomes included factors in patients with malignant disease that may predict a poor outcome from COVID-19 compared to patients without malignant disease, including patient demographics, tumour subtype and cancer treatments. A meta-analysis was performed using random effects model.

**Results:** 81 studies were included, totalling 61,532 cancer patients. Haematological malignancies comprised 22.1% (9,672 of 43,676) of cases. Relative risk (RR) of mortality when age and sex matched was 1.69 (95% CI, 1.46-1.95; p<0.001; I^2^=51%). RR of mortality, versus non-cancer patients, was associated with decreasing age (exp(b)0.96; 95% CI, 0.922–0.994; p=0.028) but not male sex (exp(b)1.89; 95% CI, 0.222–6.366; p=0.83). RR of mortality in those with haematological malignancies versus non-cancer control was 1.81 (95% CI, 1.53-2.95; I^2^=0.0%). Compared to other cancers, increased risk of death was seen for lung (RR 1.68, 95% CI, 1.45-1.94; p<0.001), genitourinary (RR 1.11; 95% CI, 1.00-1.24; p=0.059) and haematological malignancies (RR 1.42; 95% CI, 1.31-1.54; p<0.001). Breast (RR 0.51; 95% CI, 0.36-0.71; p<0.001) and gynaecological cancers (RR 0.76; 95% CI, 0.62-0.93; p=0.009) had lower risk of death. Receipt of chemotherapy had greatest overall pooled mortality risk of 30% (95% CI, 25-36%; I^2^=86.97%) and endocrine therapy the lowest at 11% (95% CI, 6-16%; I^2^=70.7%).

**Conclusions:** Cancer patients, particularly younger cancer patients, appear at increased risk of mortality from COVID-19 compared to non-cancer patients. Differences in outcomes were seen based on tumour types and treatment.

**Highlights:**

- To our knowledge this is the largest review and meta-analysis of COVID-19 in cancer patients with insights into tumour types and therapies.
- In unadjusted analysis cancer doubles the risk of COVID-19 related mortality. This decreased when adjusted for age and sex.
- Younger cancer patients have the highest risk of mortality when compared to non-cancer COVID-19 patient of a similar age.
- Patients with lung, genitourinary and haematological malignancies are at increased risk of mortality, breast and gynaecological cancers are at lower risk.
- Patients on chemotherapy have the highest pooled mortality risk with those on endocrine therapy the lowest.

## Introduction

Individuals with cancer are more prone to respiratory viruses as a result of immunosuppression from either the underlying disease or therapy. This has previously been demonstrated with influenza, which is associated with an increased mortality rate in patients with solid and haematological malignancies ^1,2^. While rhinovirus alone, if present before hematopoietic cell transplant, is associated with substantial increase in mortality ^3^. Given this, and since the emergence of the SARS coronavirus 2 (SARS-CoV-2) pandemic, there has been an intense and focused global effort to understand the impact on cancer patients of infection with SARS-CoV-2 and the outcomes with COVID-19.

Understanding the possible risks, consequences, and complications of SARS-CoV-2/COVID-19 in patients with cancer is important for patients, family, and health care systems. For patients and their families such information is key to enable informed decisions around the potential risks of anti-cancer treatment undertaken during the pandemic as well as the degree they should limit social and familial interactions. While for health-care systems, these data are vital to enable informed decisions regarding treatment risk, how to protect cancer patients as well as how best to prioritise a heterogenous population, based on cancer type and treatment, for preventive measures and anti-viral treatments. Such information is also vital in planning the response to future pandemics.

The first large population level data regarding outcome of patients with COVID-19 came from data from the United Kingdom’s pandemic protocol, International Severe Acute Respiratory and Emerging Infections Consortium (ISARIC) WHO Clinical Characterisation Protocol UK (CCP-UK). This revealed that 10% of the 20,133 hospitalised patients with COVID-19, as of 3^rd^ May 2020, had a history of malignancy. A significant increase in hospital mortality was reported in those patients with a history of malignancy (HR 1.13; 95% CI, 1.02-1.24; p=0.017) ^4^. While population level data from the Intensive Care National Audit and Research Centre (ICNARC) has found a lower proportion of patients with cancer admitted to intensive care as compared to a retrospective cohort of patients with other viral pneumonias (non-COVID-19), 2.5% versus 5.8% respectively ^5^. As well as immunosuppressed patients having increased likelihood of death if admitted to critical care ^6^.

Numerous cancer-specific studies have been undertaken as part of the effort to understand COVID-19 in cancer patients. These have found cancer patients and to have a more severe disease course, with older cancer patients ^7–27^, and those with haematological malignancies reported to be at particular increased risk ^7,8,11,23,28–33^. However, many of these studies report disparate results, particularly regarding the association of cancer type and recent cancer treatment with outcomes ^10,15,23,34,35^. These studies varied in size, nature and were limited by a lack of, or only a very small comparator non-cancer groups ^12,14,16,18,19,21,23,26,29,36–45^ as well as selectivity in which tumour types of cancer therapies were included in their analysis ^9,11,18,23,25,29,32,40,43,46–59^. The lack of a contemporaneous age and sex matched non-cancer population, variability in data collection and reporting as well as variable follow up times also limits the published cohort studies. In those studies where a non-cancer cohort was utilised, these were historical patients or based on registry data ^36^. Given all these factors, confounding biases may be present as a result of unmeasured confounders.

In the current study we undertook a systematic and unbiased review and meta-analysis of all the available published data to investigate the outcomes of patients with SARS-CoV-2 as well as identifying factors which may predict for a poor outcome. Such research and information is key to further our understanding of the impact of the SARS-CoV-2 pandemic as well as identifying possible novel risk factors in cancer patients not identified in previous cohort studies.

## Methods

### Search strategy and literature search

We undertook a systematic review of the published literature and reported our findings according to the Preferred Reporting Items for Systematic Reviews and Meta-analyses (PRISMA) guidelines ^60^. Repeated searches of PubMed, Web of Science and Scopus databases were performed up to 14^th^ June 2021. References from all identified articles were also reviewed to check for other relevant studies with duplicates identified and removed. Search strategies and results from the literature search are shown in Tables S1-S3.

### Study selection

Two authors (E.K. and C.P.) independently screened all titles and abstracts after initial de-duplication. The inclusion criteria were any case-control or cohort study with or without a control non-cancer COVID-19 group published in English as a full text article involving patients with cancer and confirmed or suspected SARS-CoV-2 infection or COVID-19, which describe one or more of: the incidence, presentation, management, or outcome of cancer patients with COVID-19. We excluded studies with fewer than 10 patients, conference papers or abstracts, preprints, articles where the full text could not be extracted, studies where data could not be obtained by contacting the corresponding author and animal studies. Where studies with overlapping datasets were present, the paper with the largest and most up to date cohorts were included. Discrepancies were resolved by consensus.

### Data extraction and quality assessment

Two authors (E.K. and C.P.) independently undertook data extraction for the following: first author, study type, time period of data collection, country of data collection, number of male and female patients, median or mean age, cancer treatment intervals prior to COVID-19 diagnosis or hospitalisation, unadjusted and adjusted odds ratios (ORs) or hazard ratios (HRs) for severe disease and death for each cancer subtype and for each cancer treatment subtype as well as the number of cancer patients with COVID-19 and cancer type. We collected data on seven cancer subtypes; breast, gynaecological, genitourinary, skin, gastrointestinal, lung and haematological malignancies, as outcome data for these subtypes was most frequently reported within our pool of 81 studies. Data was collected on participant characteristics, global distribution of the studies, setting of care and presenting symptoms.

The quality of the included studies was assessed using the Newcastle-Ottawa scale (NOS) for case control and cohort studies (Figure S1) ^61^. The eight questions within this scale were arranged into three groups: selection, comparability, and outcomes. Studies can earn a maximum of four stars from selection, two form comparability and three from outcomes, with a maximum score of 9 stars.

### Outcomes and statistical analysis

The main outcome of interest was mortality. We performed meta-analysis comparing mortality in cancer patients with COVID-19 to control patients (with COVID-19 but without cancer) and in cancer patients with each tumour subtype compared to cancer patients who did not have that subtype. We also performed meta-analysis pooling case fatality rates for patients with each tumour subtype and those who had received each type of cancer treatments.

For meta-analysis of mortality comparing cancer patients and controls and comparing cancer patients with and without tumour subtypes, results are presented as pooled risk ratio (RR) and 95% confidence intervals (CIs). Meta-analyses of case fatality rate by tumour subtype and cancer treatment results are presented as pooled proportions and 95% CIs.

As statistical heterogeneity was anticipated due to expected variability in study design and participant characteristics, all meta-analyses were conducted using a random effects model, with restricted maximum likelihood to estimate between-studies heterogeneity. Statistical heterogeneity was quantified using the I^2^ statistic. To examine the impact of age and sex on mortality among cancer patients and controls, random-effects meta-regressions were conducted. Meta-analysis and meta-regression were conducted in Stata version 14.1 using commands admetan ^62^, metaprop ^63^ and metareg ^64^. Additional supplementary figures (of study characteristics were produced in R version 4.0.4 (Figures 4, Figure S3 – 6, S9, S11 & S14a-b).

## Results

The initial search retrieved 1,150 articles for review (Figure S2). After the inclusion of records identified through additional sources and removal of duplicate articles, 1,004 records were screened. We obtained 215 articles to assess for eligibility. Of these, 134 were excluded (see Figure S2 for reasons). Eighty-one studies were included in this systematic review and meta-analysis.

### Global distribution of studies

Eighty-one studies were included in this systematic review, with a total of 61,532 cancer patients with COVID-19 (Figure S3, Table S4). 61 were retrospective ^7,8,10–14,16–25,29–32,36–45,48,49,53–55,57–59,65–85^, 17 were prospective ^4,26–28,33,35,46,50–52,56,86–91^ and three were retro-prospective studies, whereby data were collected on both patients eligible prior to study commencement and those who enter the study post study commencement ^9,15,47^. Patients were recruited across 28 countries and 5 continents (Figure 1, Figure S4, Table S5). Of the 80 studies that provided information on recruitment by country, the five countries contributing the most patients were: USA, United Kingdom, Italy, France, and China.

**Figure 1.**
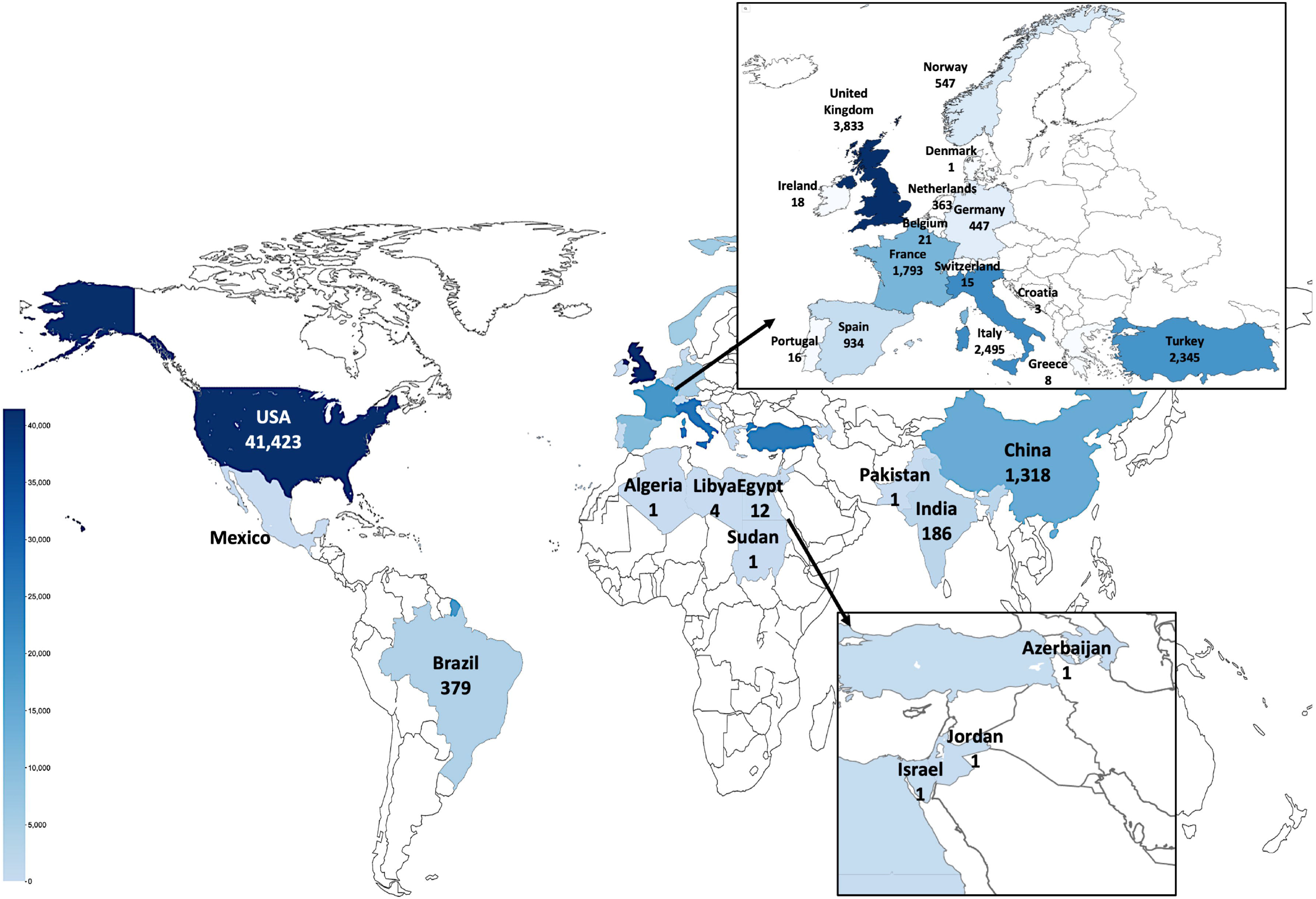
Distribution of patients across 80 studies.

### Patient characteristics and cancer types

Ten of 81 studies (12%) included cancer patients amongst a cohort that included non-cancer patients with COVID-19 ^4,44,65,72,75,79,80,84,86,89,91^; however, the majority of studies (n=71, 88%) reported on cohorts of cancer patients only. The number of cancer patients in the 81 reports ranged from 11 to 38,614. Where data were available, 52% (30,557 of 58,849) were male, age ranges from 10 to 98 years (median age range 35 to 74 years). The most frequent comorbid conditions reported were hypertension, diabetes mellitus, cardiovascular and pulmonary disease (Figures S5 – S7 & S10b and c). The majority (34,117; 55%) were hospitalised patients and the remainder were outpatients or unknown (Figure S9 and Table S7).

The majority of reports (n=55, 68%) included patients with both solid and haematological malignancies (total of 55,668 patients). 18 (22%) reported on patients with haematological malignancies alone (n=2,526 patients) ^11,17,18,23,25,29,32,40,43,50,51,53,54,56,58,59,67,77^. Of these 18 studies: four reported exclusively on multiple myeloma and two on chronic lymphocytic leukaemia (CLL) patients. While 8 (10%) studies reported exclusively on patients with solid malignancies ^9,46–49,52,55,57^, of these, 2 studies each reported on thoracic cancers ^46,49^, gynaecological cancers ^48,57^ and breast cancer ^52,55^. Tumour type was reported in 68 studies (84%) for 43,676 patients (71%) (Table S6). The most frequent were haematological malignancies representing 9,672 (22.1%) of patients followed by breast 8,322 (19.1%); genito-urinary 7,624 (17.4%); skin and melanoma 6,613 (14.1%); gastrointestinal 4,124 (9.4%) and thoracic cancer 2,104 (4.8%). For 1,139 patients (2.6%), the tumour type was documented as other or unknown (Figures S8 & S10A).

### Presenting symptoms of SARS-CoV-2 infection

Fifty-three (65%) of the 81 studies, involving 9,196 patients, reported presenting symptoms. The most commonly reported symptoms were fever, cough, dyspnoea, fatigue/malaise, diarrhoea, and myalgia (Figures S10 & S11). The time from onset of symptoms to hospitalisation ranged between 1-40 days, where reported ^10,46,48,53,66,69,78^.

### Radiological findings

Radiological findings of thoracic imaging based on chest radiograph, CT scanning or a combination of chest radiograph and CT scan in cancer patients with COVID-19 were available in 28 studies (n=3,650 patients) ^10,13,14,17,25,31–33,35,39,42,43,45,50,52,54–58,66,69,78,81,83–85,87^. A total of 19 (26%) studies (n=3,086 patients) provided details of radiological changes (Table S8) ^10,13,14,25,31–33,35,39,42,43,50,52,54,56,66,78,85,87^. CT features of ground glass opacity, patchy shadows, fibrous stripes, pleural thickening, bilateral involvement, and tumour pulmonary involvement in cancer patients were associated with worsening severity of COVID-19 disease ^14,35,66,78,85,87^.

### Mortality of cancer patients compared with controls

Nineteen of 81 studies (23.5%), compared patients with cancer and COVID-19 to a cohort of patients without cancer and COVID-19 ^12,14,16,18,19,21,23,26,29,36–43,45^. The details of these studies, the matching of the cohorts and the nature of the cohort without cancer and COVID-19 are described in Table 1.

**Table 1.** Comparison of malignant versus non-malignant patients with outcomes in 19 studies. * Denotes studies looking at patients with haematological malignancies.

| Publication | Comparison | Cancer patients | Mortality | Non-cancer patients | Mortality | Features |
| --- | --- | --- | --- | --- | --- | --- |
| Yigenoglu et al, 2020 * | Non-cancer COVID-19 patients (age, gender and comorbidity matched) | 740 | 102 | 740 | 50 | Patient characteristics and outcomes |
| Johannesen et al, 2021 | Non-cancer COVID-19 patients | 547 | 56 | 7,841 | 158 | Patient characteristics and outcomes |
| Rüthrich et al, 2020 | Non-cancer COVID-19 patients (age matched) | 435 | 97 | 2,636 | 367 | Patient characteristics and outcomes |
| Montopoli et al, 2020 | Non-cancer COVID-19 patients | 430 | 75 | 4,532 | 313 | Patient characteristics and outcomes |
| Miyashita et al, 2020 | Non-cancer COVID-19 patients (age matched) | 334 | 37 | 5,354 | 518 | Patient outcomes |
| Lunski et al, 2020 | Non-cancer COVID-19 control cohort (Ochsner Health System) | 157 | 56 | 1,460 | 372 | Patient characteristics laboratory markers, and outcomes |
| Tian et al, 2020 | Non-cancer COVID-19 patients (1:2 Propensity score matched) | 232 | 46 | 519 | 56 | Patient characteristics laboratory markers, and outcomes |
| Mehta V et al, 2020 | Non-cancer COVID-19 patients, 1:5 Propensity matched (age and gender matched) | 218 | 61 | 1,090 | 149 | Patient characteristics and outcomes |
| Martinez-Lopez et al, 2020 * | Non-cancer COVID-19 patients (age and gender matched) | 167 | 56 | 167 | 38 | Patient characteristics laboratory markers, and outcomes |
| Brar et al, 2020 | Non-cancer COVID-19 patients, 1:4 (age, gender, comorbidities matched) | 117 | 29 | 468 | 100 | Patient characteristics and outcomes |
| Meng et al, 2020 | Non-cancer COVID-19 patients (1:3 Propensity score matched) | 109 | 32 | 327 | 40 | Patient characteristics laboratory markers, and outcomes |
| Dai et al, 2020 | Non-cancer COVID-19 patients (age matched) | 105 | 12 | 536 | 21 | Patient characteristics and outcomes |
| Cattaneo et al, 2020 * | Non-cancer COVID-19 patients (matched by age, gender, comorbidities and respiratory failure) | 102 | 40 | 102 | 24 | Patient characteristics and outcomes |
| Shah et al, 2020 * | Non-cancer COVID-19 patients (age and gender matched) | 80 | 31 | 1,115 | 223 | Patient characteristics and outcomes |
| Sun et al, 2021 | Non-cancer COVID-19 patients | 67 | 9 | 356 | 4 | Patient characteristics and outcomes |
| Joharatnam-Hogan et al, 2020 | Non-cancer COVID-19 patients (matched by age, gender, comorbidities) | 30 | 11 | 90 | 32 | Patient characteristics laboratory markers, and outcomes |
| Stroppa et al, 2020 | Non-cancer COVID-19 patients (matched by age, gender, pneumonia and antiviral treatment) | 25 | 9 | 31 | 5 | Patient characteristics and outcomes |
| Liang et al, 2020 | Non-cancer COVID-19 patients | 18 | 7 | 1,572 | 124 | Patient characteristics and outcomes |
| He et al, 2020 * | Non-cancer COVID-19 health care providers | 13 | 8 | 11 | 0 | Patient characteristics laboratory markers, and outcomes |

We conducted a meta-analysis of these 19 studies. The pooled relative risk (RR) of mortality in cancer patients with COVID-19 compared with non-cancer control patients with COVID-19 was 2.12 (95% CI, 1.71-2.62, p<0.001, I^2^ = 84.4%, Figure 2a). When pooling results, the relative risk for mortality in 13 studies that matched for age was reduced to RR 1.69 (95% CI, 1.46-1.95; I^2^ = 51%; p<0.001) compared to RR 3.80 (95% CI, 2.53-5.71; I^2^=81.9%, p<0.001) from six studies without matching (Figure 2b). There was little difference in the pooled RR of mortality between any type of malignancy, solid or haematological, (RR 2.23; 95% CI, 1.68-2.95; I^2^=88.7%) and haematological malignancies (RR 1.81; 95% CI, 1.53-2.95; I^2^=0.0%) alone compared to non-cancer control patients, respectively (Figure S12).

**Figure 3.**
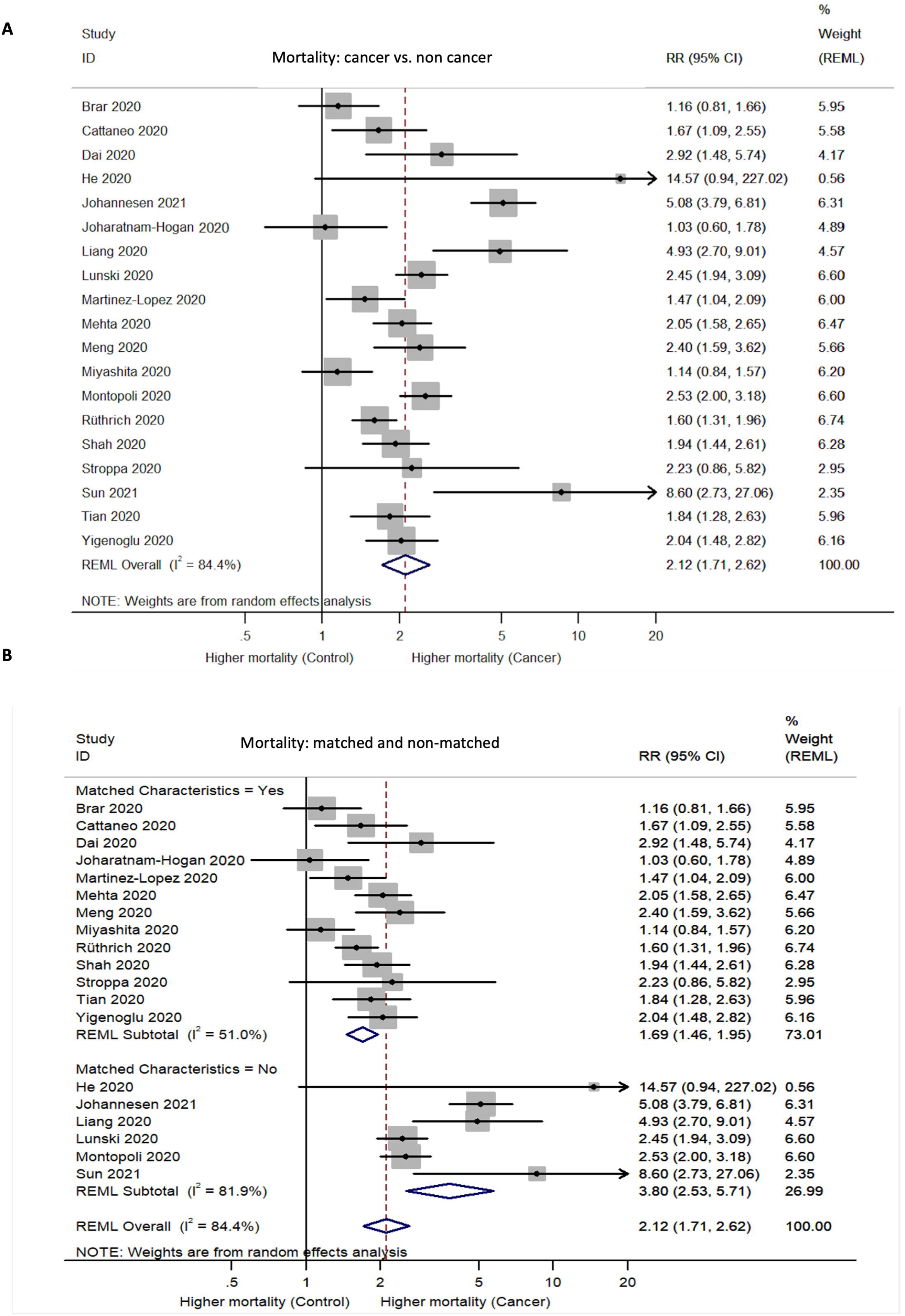
Forest plot of relative risk of mortality in analysis. A) Unadjusted mortality risk due to presence of malignant disease. B) Relative risk of mortality due to the presence of malignant disease, after matching for age and gender.

Fourteen studies provided data on the median or mean age of cancer patients and non-cancer patients with COVID-19 ^14,16,18,19,21,23,26,29,39–43,45^. On univariate regression, when assessing the impact of age on the comparison of cancer and control patients, the RR of mortality statistically significantly decreased as age increased (exp (b) 0.96; 95% CI, 0.922 – 0.994; p = 0.028), shows a greater difference in the relative risk of mortality between cancer patients and controls of younger ages (Figure 3a). Seventeen studies reported the sex (as proportion of males) of both cancer and non-cancer control groups ^14,16,18,19,21,23,26,29,37–45^. A small increase in RR of mortality between cancer patients and controls was shown as the proportion of males in the study increased, but this was not statistically significant (exp (b) 1.89; 95% CI, 0.222–6.366; p=0.83; Figure 3b). When the combined impact of age and proportion of males was explored by multivariable regression, age remained significant (p=0.029) and male sex not significant (p=0.545) (Table S9).

**Figure 3.**
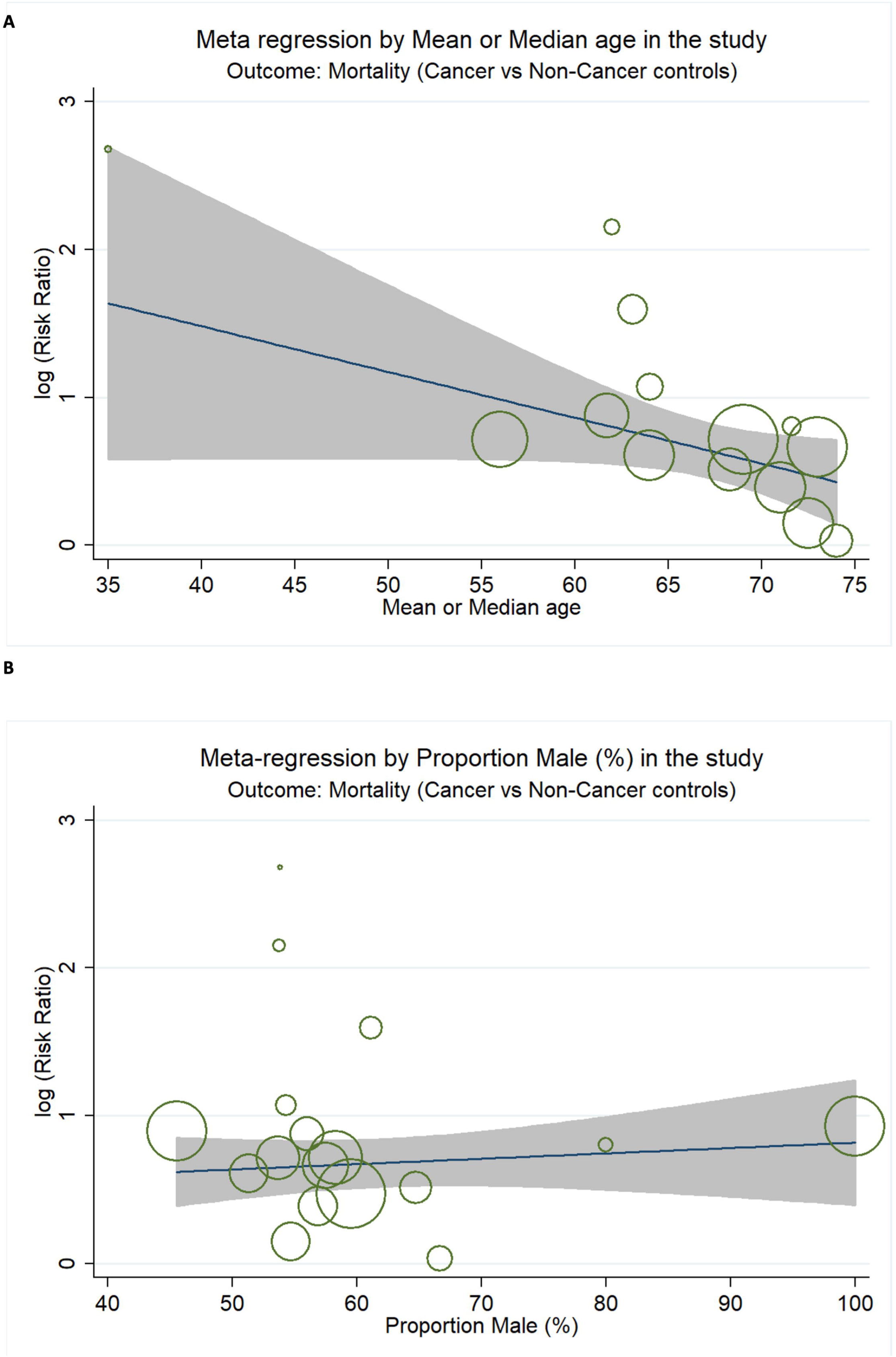
Meta-regression bubble plot results of impact of age and gender. (A) Risk ratio of mean or median age in cancer vs non cancer subgroups. (B) Risk ratio of male

### Clinical outcomes

Outcome data were available for 56,932 patients with cancer and COVID-19 from 68 (84%) of the 81 studies ^7–29,31–59,66–70,74,75,77,78,81–84,87,89,90^. Where reported, 7.1% (1,170 of 16,409) were admitted to intensive care unit (ICU) and 5.2% (2,817 of 54,298) required invasive mechanical ventilation (IMV) (Table S10). At the time of reporting, 10.7% (473 of 4,403) of patients remained hospitalised and 64.5% (2,841 of 4,403) were discharged. Median duration of hospital stay is shown in Table S11. Of the 56,932 cancer patients, 6,813 died (12%). Median follow up times varied between 5 and 69 days. Mortality was reported in all studies and ranged from 3.7% to 61% (Figure 4, Table S12). Of note, one study defined mortality as either transfer to hospice or death ^22^.

**Figure 4.**
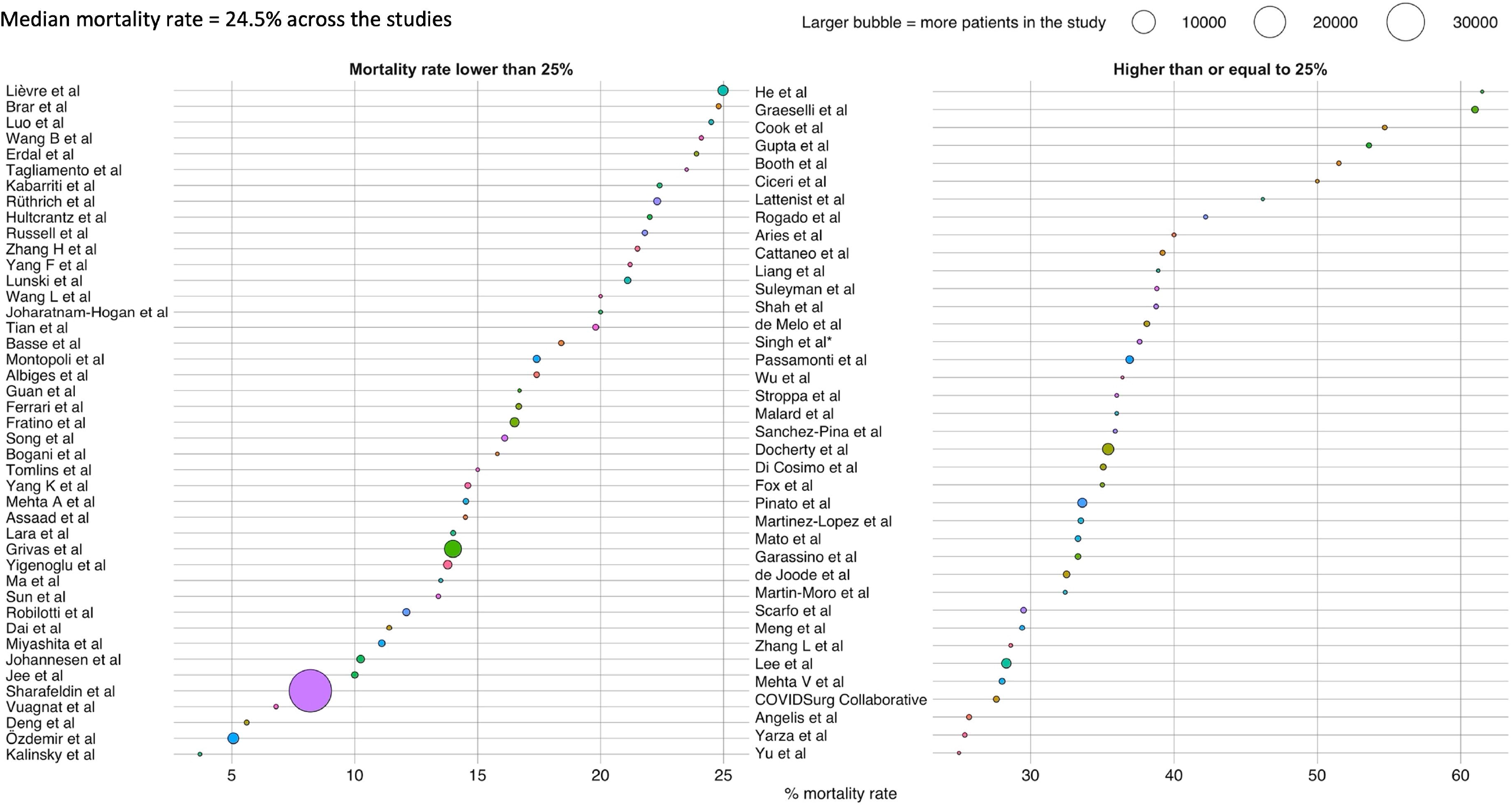
Mortality reported across 81 studies.

14 different definitions of severe events were used across the studies (Figure S13). 12 studies reported information on ethnicity and outcomes, (summarised in supplementary results document). In unadjusted analyses, several factors were found to be significantly associated with severe events or death. These included increasing age (in 36 studies), raised pro-inflammatory (in 14 studies) and infection related markers (in 14 studies), Figure S14a. Factors found to be significantly associated with worsening severity or mortality, in adjusted analysis are shown in Figure S14b, of which 22 studies found increasing age to be associated with worsening severity or death ^7–27^. Factors included in adjusted analysis are listed in Figure S15.

### Mortality and case fatality rate by cancer type

Pooled case fatality rate forest plots for each cancer subtype are shown in Figure S16. There was a 9% (95% CI, 7% to 12%, I^2^ = 89.8%; range: 0% to 100%) pooled case fatality rate for patients with breast cancer with a RR of mortality of 0.51 (95% CI, 0.36-0.71; p<0.001; I^2^=86.2%) when compared with those with non-breast cancers and COVID-19 (Figure 5a). The pooled case fatality rate of patients with COVID-19 and gynaecological cancers was 12% (95% CI, 8%-16%; I^2^ = 38.47%; range: 0% to 38%) and an associated RR of 0.76 (95% CI, 0.62-0.93; p=0.009; I^2^= 0%), between those with gynaecological cancers and patients with non-gynaecological cancers (Figure 5b).

**Figure 5.**
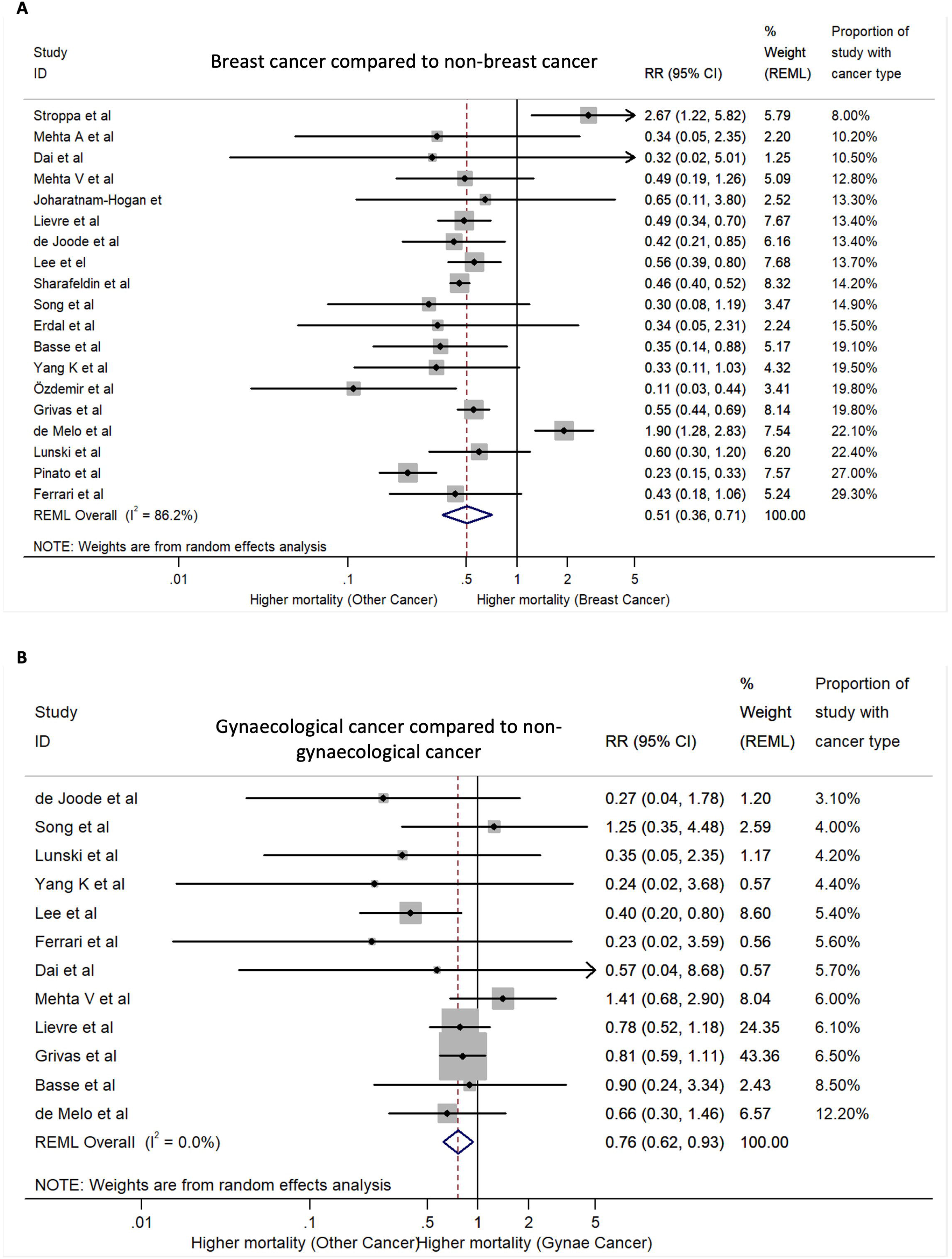
Forest plot of relative risk of mortality in the subgroup analysis. A) Breast cancer and COVID-19, compared to patients with non-breast cancers and COVID-19. B) Gynaecological cancer and COVID-19, compared to patients with non-gynaecological cancers and COVID-19.

Gastrointestinal cancers were associated with a pooled case fatality rate of 16% (95% CI, 12-20%; I^2^=78.66%; range 0% to 38%) and a RR of 1.13 (95% CI, 0.93-1.37; p=0.207; I^2^=54.8%) compared to patients with non-gastrointestinal cancers (Figure S17b). Skin cancer was associated with a pooled case fatality rate of 10% (95% CI, 5% to 15%; I^2^=62.57%; range: 5% to 50%) and a relative risk of mortality of 0.85 (95% CI, 0.60-1.20; p=0.353; I^2^=51.4%; Figure S17a).

The pooled case fatality rate of patients with COVID-19 and lung cancers was 30% (95% CI, 24%-37%; I^2^=83.71%; range: 0% to 60%). The RR of mortality in those with lung cancer compared to other cancer types was significantly higher at 1.68 (95% CI, 1.45–1.94; p<0.001; I^2^ = 32.9%, Figure 6a). Pooled case fatality rate for genitourinary cancers was 22% (95% CI, 16% to 27%; I^2^=92.61%; range: 8% to 50%), with a RR of mortality of 1.11 (95% CI, 1.00-1.24; p=0.059; I^2^ = 21.5%,) when compared to non-genitourinary cancers (Figure 6b). The pooled case fatality rate in patients with haematological malignancies was 32% (95% CI, 28%-37%; I^2^=93.10%; range: 11% to 100%). The overall relative risk of mortality in cancer patients with haematological malignancies, compared to those with solid malignancies is 1.42 (95% CI, 1.31-1.54; p<0.001; I^2^=6.8%, Figure 6c).

**Figure 6.**
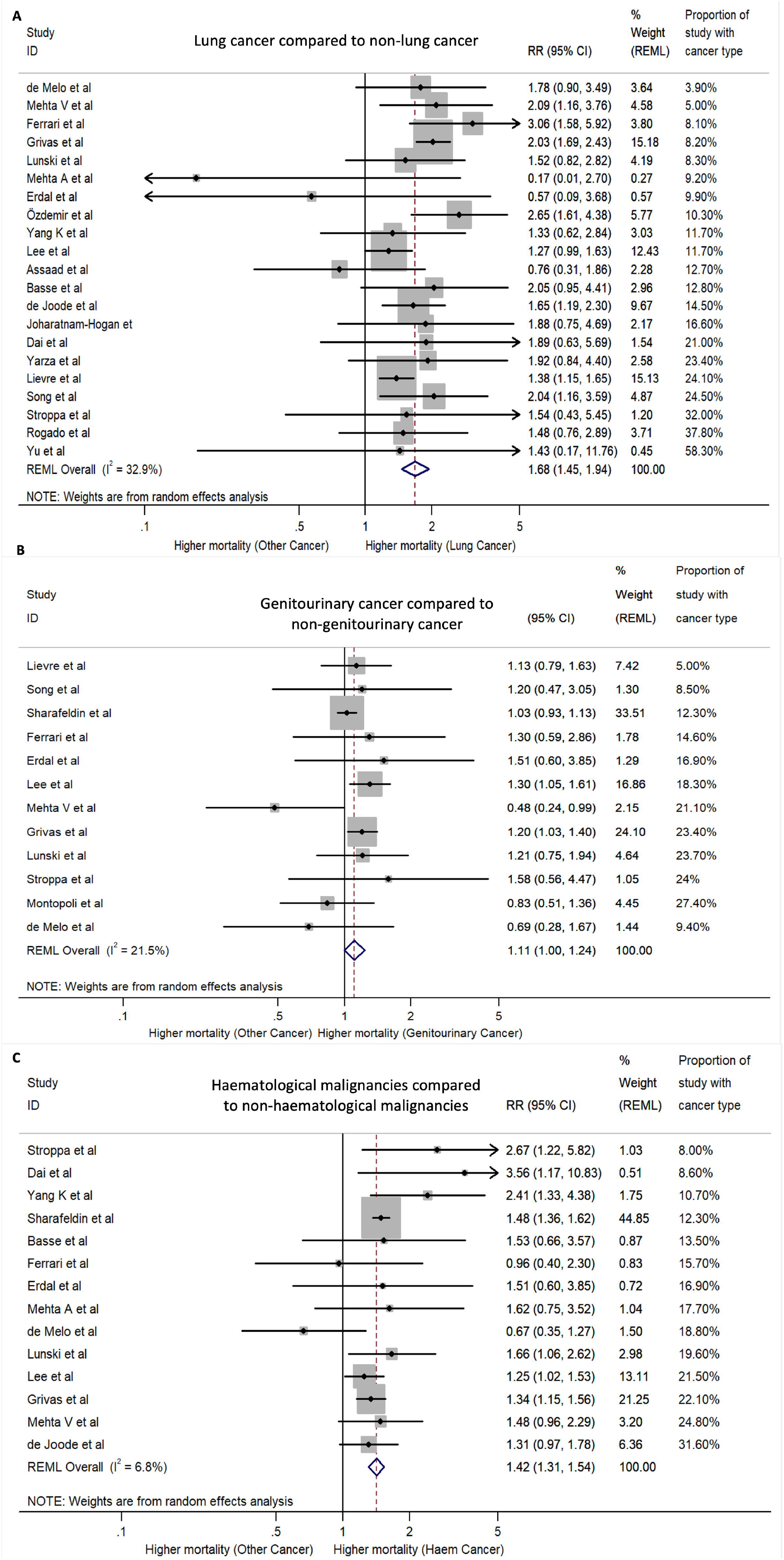
Forest plot of relative risk of mortality in subgroup analysis. Relative risk of mortality in different cancer types: A) Patients with lung cancer and COVID-19 compared to patients with non-lung cancers and COVID-19. B) Patients with genitourinary cancer and COVID-19 compared to patients with non-genitourinary cancers and COVID-19. C) Patients with haematological malignancies and COVID-19 compared to patients with non-haematological malignancies and COVID-19.

### Stage of malignancy and outcome

There was variability with regards to the reported effect of stage of malignancy on outcomes with COVID-19. In unadjusted analysis, 8 studies have reported stage IV disease (of solid cancers) to be associated with significantly higher risk of severe events and mortality compared to patients with stage I, II or III disease ^9,10,14,21,66,68,83,85^. However, five studies found no association with metastatic primary disease late stage of disease and rates of hospitalisation or mortality ^22,30,31,35,46^. A study of 181 patients in Brazil found that those with 2 or more metastatic sites had an overall increased risk of death ^26^.

### Cancer treatment and course of COVID-19

Data on timing of cancer treatment and outcome were available in 64 of 81 (79%) of the reported studies (Figure S18). These studies comprised 54,335 patients of whom 7,567 (13.9%) had undergone or received some form of treatment for their malignant disease. Where data was available, the commonest treatment modality was chemotherapy (n=3,792), followed by targeted therapy (n=1,700) and immunotherapy (n=817). 3,896 patients were reported as not having received any form of treatment.

Surgery, within 3 months of a diagnosis of COVID-19 in cancer patients was associated with a pooled case fatality of 19% (95% CI, 9%-29%; I^2^=58.49%; range: 0% to 40%, Figure S18a). These studies in general looked at all patients, aside from one which was focused on surgery for gynaecological malignancies ^48^. Patients with cancer who had received chemotherapy and were diagnosed with COVID-19 had an overall pooled case fatality of 30% (95% CI, 25%-36%; I^2^=86.97%; range: 10% to 100%, Figure S18b). While endocrine therapy had a pooled mortality risk of 11% (95% CI, 6%-16%; I^2^=70.68%; range: 0% to 27%, Figure 18c). 9 studies did not specify which cancer subtype was treated with endocrine therapy. Immunotherapy was associated with a pooled case fatality of 19% (95% CI, 16%-28%; I^2^=36.98%; range: 0% to 50%, Figure S18d). Two of these studies specified the tumour involved, lung cancer ^49^ and gynaecological cancer ^48^, with the remaining studies not specifying tumour type. Mortality following radiotherapy was available in 10 studies, with a pooled case fatality of 23% (95% CI, 12%-33%: I^2^=74.84%; range: 0% to 50%, Figure S18e). Tumour type treated, gynaecological, was specified in only one study ^48^. Targeted therapy was associated with an overall risk of mortality of 18% (95% CI, 12%-23%; I^2^=59.72%; range: 0% to 57%, Figure S18f). The nature of the targeted therapy was not specified in most cases and the specific cancer type was only stated in 4 of 19 studies (21%): two involving patients with haematological malignancies ^25,58^, and one respectively for patients with lung ^49^, and gynaecological cancer and COVID-19 ^48^. These data are in Table S13.

## Discussion

From the start of the SARS-CoV-2 pandemic, there has been a focus to identify the patients at greatest risk of poor outcome from infection. Patients with malignant disease were rapidly identified as one such group ^92^. Initial reports of SARS-CoV-2 infection in cancer patients were small and retrospective ^26,43,57,58,81,82,90^, subsequently larger collaborative registry studies, prospective and retrospective, have been reported ^4,7,37,46,47,93,94^. These studies have varied in geographical location and focus with regards to tumour type (Table S4). This systematic review and meta-analysis sought to analyse in an unbiased fashion this important global effort by the cancer community by reviewing all of the available and published data as of the June 14^th^ 2021. The aim being to understand the impact of cancer type, stage and treatment on outcomes from COVID-19 as well as identifying possible novel risk factors in cancer patients not identified in previous studies.

This systematic review demonstrates a number of potential limitations with the currently available data and literature these include; (1) lack of contemporaneous non-cancer populations for comparative analysis ^12,19,23,36,38,40^; (2) heterogeneity of definitions between studies such as severity of COVID-19 as demonstrated by the 14 definitions of severity used across studies (Figure S13;) (3) predominantly retrospective nature of studies (Table S4, 75%); (4) variable follow up; (5) heterogeneity or poor description of the non-cancer control cohorts, such as use of non-hospitalised patients ^12,21,29,36–38,44^, or staff members with COVID-19 ^43^; and (6) lack of detail relating to systemic cancer treatment ^4,11,12,22,29,33,37,39,41,42,50,52,55–57,65,70–72,75,76,79–81,84,86–91^. In those studies where a non-cancer cohort was utilised these data were generally historical or based on registry data and were not contemporaneous to the cancer cohort ^12,19,36,38,43^. Only three of the 19 studies that compared cancer cohort to a matched non-cancer cohort used propensity score matching ^14,16,39^. Therefore, within the published literature biases may be present as a result of unmeasured confounders.

Utilising data from 19 studies with 3,926 patients with 38,847 non-contemporaneous non-cancer patients, we found that malignant disease is associated with an increased risk of severe or death from COVID-19 compared with non-cancer patients (RR 2.12; 95% CI, 1.76-2.62; p<0.001; I^2^=84.4%). In particular, we demonstrate via regression analysis that younger age is associated with a worse clinical outcome relative to age matched non-cancer patients. To date, all the cohort studies, which by their nature have lacked an age matched control, have consistently reported increasing age as a risk factor for poor clinical outcome ^7–27^. While it is true older patients have a worse absolute outcome than younger patients, the relative risk, as shown in this review, is highest for the younger patients. This observation has been reported within the International Severe Acute Respiratory and emerging Infections Consortium (ISARIAC) WHO Coronavirus Clinical Characterisation Consortium (CCP-Cancer UK) ^93,94^. In the most recently presented analysis of over 20,000 cancer patients compared with 155,000 non-cancer patients, patients aged less than 50, particularly on active treatment, were five times more likely to die than similar aged non-cancer patients (<50 years, HR 5.22; 95% CI, 4.19 to 6.52; p <0.001). When compared with non-cancer patients the relative risk of death (the cancer attributable risk) fell with age ^94^. The reasons for this are likely to be related to the type of cancers, more intensive treatments, or possibly behavioural factors such as increased social mixing as compared with an older population.

Our meta-analysis also demonstrates that cancer patients with COVID-19 are twice more likely to die compared to non-cancer matched cohorts. However, when we matched these patients for age and sex, the risk decreased to 1.69 (95% CI 1.46-1.95, p<0.001; I^2^=51%). This risk, while significant, is lower than that reported by studies that did not match for these factors, where the risk was nearly four times greater in the cancer cohort ^26,36,38,41,43,44^. This demonstrates that there has been a potential overestimation of the true risk to cancer patients in studies that did not adjust for these factors. This also highlights the critical importance of a comparator non-cancer cohort in understanding the true effects of SARS-CoV-2 within the cancer population. We did not observe any significant differences in outcome by sex when cancer and non-cancer patients were compared again, this is different from a number of studies which have reported male sex as a risk factor ^7,10,18,22,23,28,31,66^.

Our study found that patients with lung cancer, followed by haematological malignancies, were at greatest risk of mortality from COVID-19, when compared with patients with other cancers. Haematological malignancies have been consistently reported as being a risk factor for poor outcome from COVID-19 ^7,8,11,23,28,31,32^ and the current meta-analysis demonstrates that such patients are 1.42 times (95% CI, 1.31-1.54; p <0.001; I^2^=6.8%) more likely to die compared with other cancer patients and 1.8 times (95% CI, 1.53-2.95; I^2^=0.0%) more likely to die compared with non-cancer COVID-19 patients. The increased susceptibility of patients with haematological malignancy is in keeping with the more profound immune suppression that affects this patient group. The increased in mortality with lung cancer (RR 1.68; 95% CI, 1.45–1.94; p<0.001; I^2^ = 32.9%) compared with patients without lung cancer is likely a reflection of age, the reduced lung reserve, associated co-morbidities associated, and possible effects of cancer treatment. The reason for the lower mortality from COVID-19 seen in patients with breast and gynaecological malignancies is not clear. It could be related to the protective effect of the female sex although we find no difference in outcomes based on sex with our meta-analysis. A possible alternative explanation relates to the hormonal milieu in these patients, with patients with both breast and gynaecological cancers have low circulating oestradiol levels. Breast cancer patients by virtue of the fact they are generally post-menopausal and often on aromatase inhibitors. While treatment for gynaecological cancers often necessitate removal of the ovaries. The possible influence of endocrine therapy has been reported for patients with prostate cancer where the use of androgen deprivation therapy has been associated with a protective effect from SARS-CoV-2 infection ^38^. There is an ongoing need to dissect the impact of female sex hormones on infection with SARS-CoV-2 as well as the course of COVID-19.

With regard to cancer treatments our pooled case fatality analysis found endocrine therapy to have the lowest pooled case fatality rate at 11% with chemotherapy having the highest at 30%. The higher mortality seen with chemotherapy compared to other treatments is likely reflective of the immunosuppressive effects chemotherapy. Cohort studies have reported disparate findings with regard to the risk of chemotherapy, with some finding significant increase risk of death ^8,9,26,28,36,40,46,66,85^, and others finding no association ^13–16,20,24,25,27,30,31,34,35,48,58,68,74^. The lack of patient level data means we have been unable to define the risk of mortality for cancer patients receiving anti-cancer therapy who develop COVID-19. Similarly, for a comparison of the impact of cancer treatment on the risk of mortality when compared to non-cancer patients. A comparison of risk by different treatment modalities as well as by individual agents also was not possible given it was not clear if patients had received more than one treatment modality and individual drugs were not named. For a more informed approach and discussion regarding the risk of different anti-cancer therapy in the context of COVID-19 will require more granular information as well as a comparison to a non-cancer population controlled by age and sex.

This systematic review and meta-analysis has a number of limitations given the nature of the data published and available for analysis. Firstly, we assessed outcomes in 7 tumour types as outcome data for these were most frequently reported within the reported studies, thus we were not able to examine the relative risk of mortality of other cancer types. Secondly, it was not possible to compare solid cancers to a control non-cancer cohort as there were no suitable studies identified. Thirdly, we are unable to explore the potential effects of different SARS-CoV-2 variants given this information was not available. Finally, the data included in this meta-analysis is in the pre-vaccination and anti-viral medication part of the pandemic, and therefore vaccinations and active treatments may impact on the observations made within this meta-analysis.

## Conclusion

In this large and comprehensive analysis of the available data we demonstrate a higher risk of death from COVID-19 for cancer patients when compared to non-cancer patients, although the risk is lower than that reported by individual cohort studies. Younger patients are shown to be at particular increased risk of poor clinical outcomes when compared to age-matched controls. With lung cancer patients having the highest risk of mortality, along with patients with haematological malignancies. Given these data, younger patients must be classified as being a high-risk population. On-going studies such as CCP-Cancer-UK will enable a comparison of non-cancer versus cancer patients controlled for age, sex and other co-morbidities as well as dissecting the true risk different tumour types and treatment controlling for patient level factors. Studies are also needed to assess the impact of mitigation and treatment measures over the time course of the pandemic between cancer and non-cancer patients particularly given many of the treatment studies did not recruit cancer patients. The global effort to understand the effects of SARS-CoV-2 on cancer patients has resulted in a rich data resource which should be utilised for an individual patient level meta-analysis. Such data will be vital in ensuring we maximize our learning and knowledge and to prepare the cancer community for the subsequent pandemics which will inevitability occur.

## Supporting information

Supplementary Results

## Data Availability

All data produced in the present study are available upon reasonable request to the authors.

## Acknowledgments

The authors acknowledge support from UK Research Innovation (UKRI)-Department for Health and Social Care (DHSC) COVID-19 Rapid Response Rolling Call (Grant reference MR/V028979/1), The Liverpool Experimental Cancer Medicine Centre [Grant Reference: C18616/A25153], Cancer Research UK, The Clatterbridge Cancer Charity and North West Cancer.

