## Supplementary Results for "A comprehensive systematic review and meta-analysis of the global data involving 61,532 cancer patients with SARS-CoV-2 infection"

Ethnicity

12 studies looked at association of ethnicity in cancer patients and outcomes, of which 8 found that African-American patients had more severe COVID-19 and increased mortality compared to white patients on both unadjusted analysis ^1-5^ and multivariable anaylsis^6-8^. Two studies found non-white patients to be significantly associated with hospitalisation and severe illness compared to white race ^14,40^; and Asian ethnicity was associated with increased mortality [HR 3.73; 95% CI: 1.28-10.91] in an adjusted analysis ^28^ While 10 studies did not find a significant association between ethnicity and poor outcome^6,9-16^. One of these studies found that hospitalised black cancer patients had lower mortality rates in adjusted analysis (OR 0.72, 95% CI 0.53-0.98) compared to non-white Hispanic patients ^17^. Hispanic patients did not have increased risk of mortality compared to non-Hispanic patients ^16^. Non-white race was found to be significantly associated with hospitalisation and severe illness compared to white race ^5,7^. Asian ethnicity was associated with increased mortality [HR 3.73; 95% CI: 1.28-10.91] in adjusted analysis ^18^.
