## Supplementary figures and images for "A comprehensive systematic review and meta-analysis of the global data involving 61,532 cancer patients with SARS-CoV-2 infection"

### Figure S1

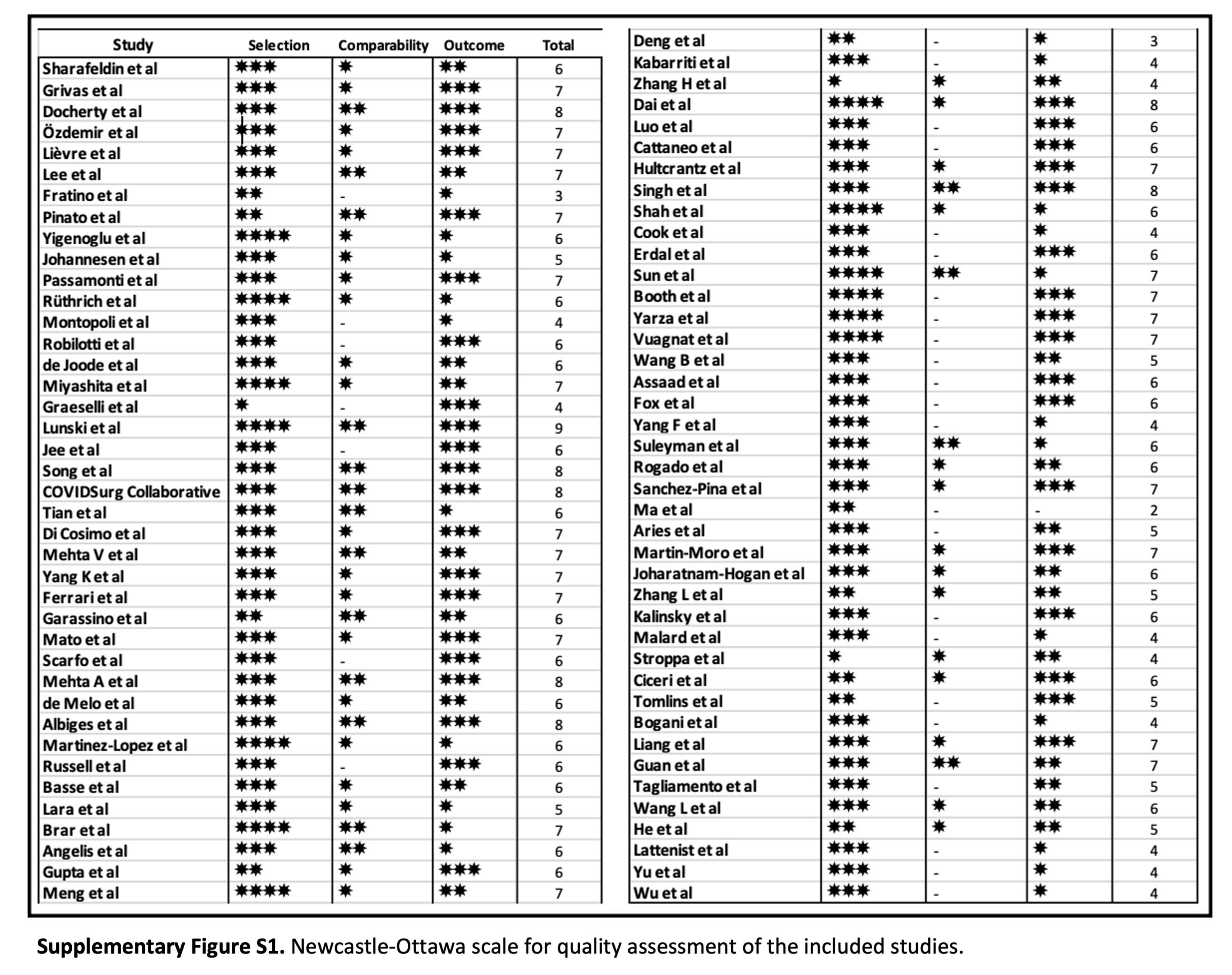

### Figure S2

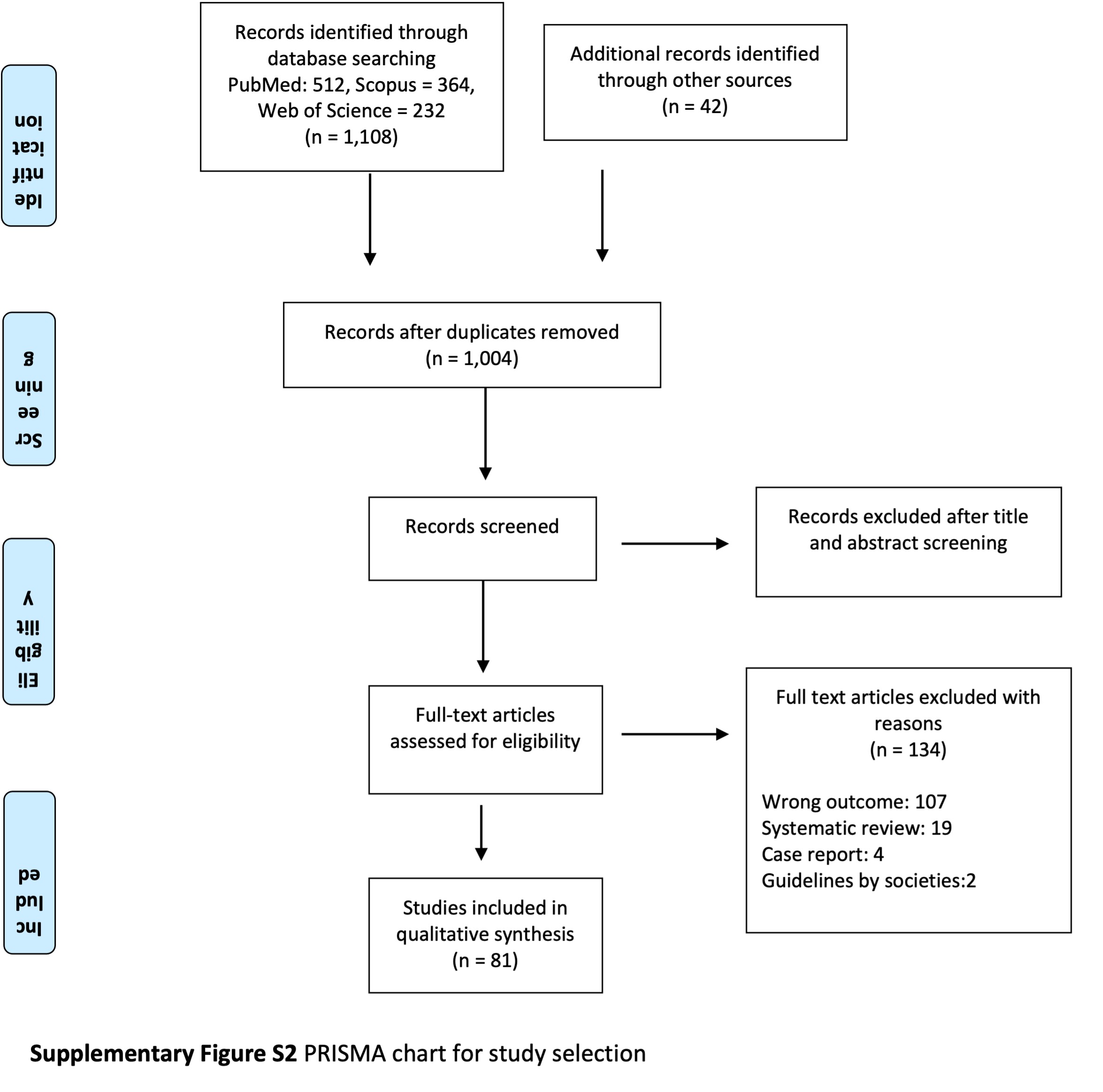

### Figure S3

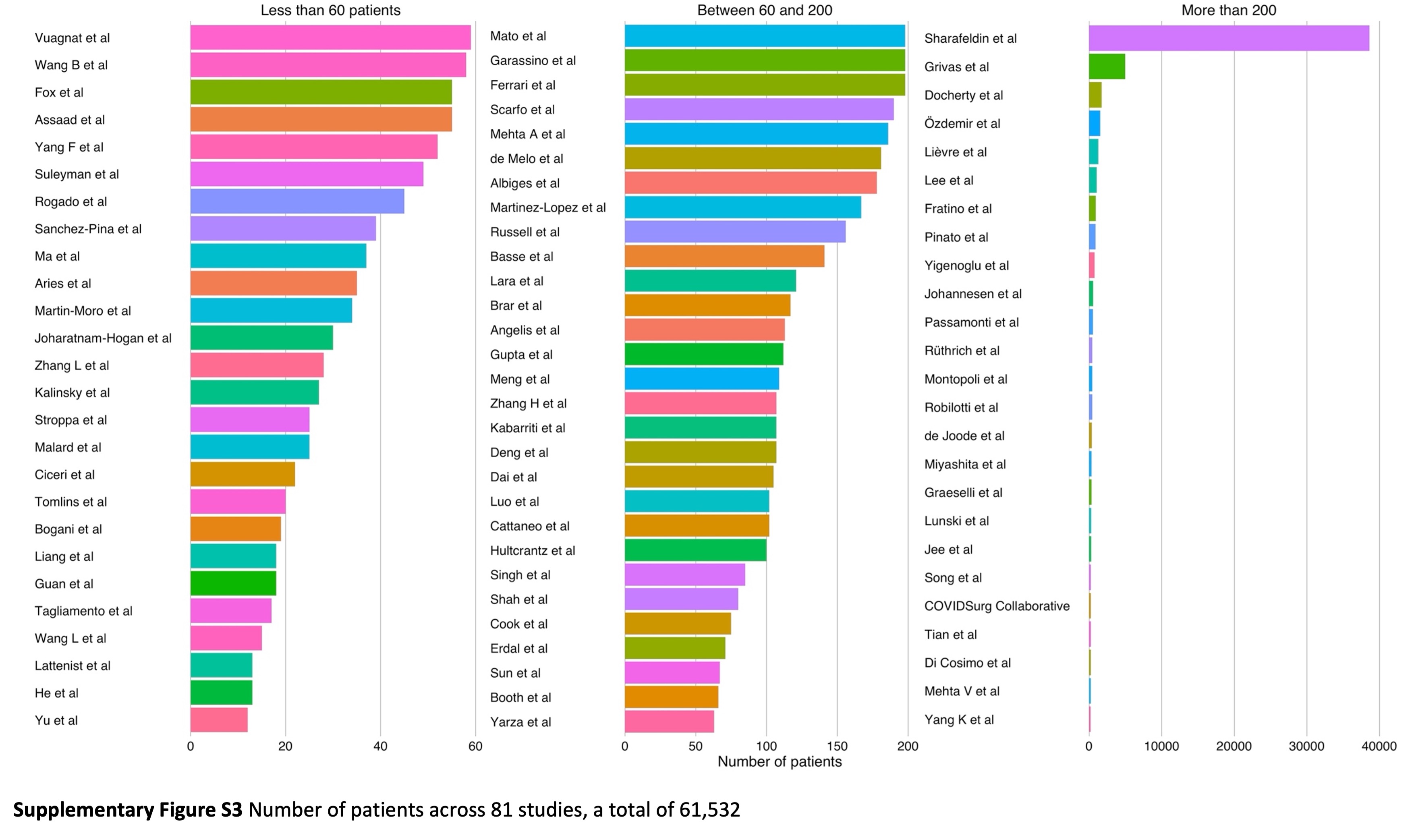

### Figure S4

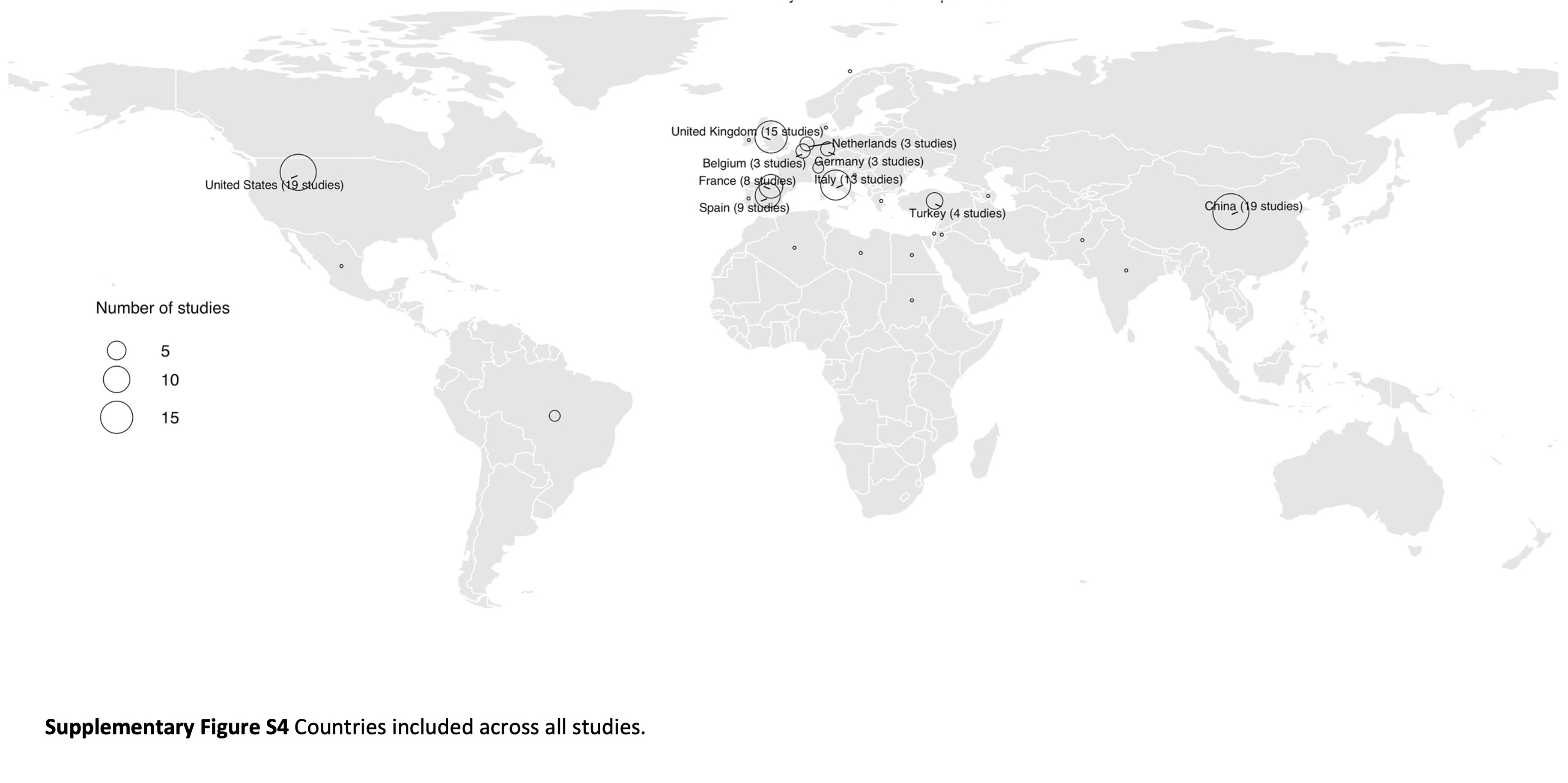

### Figure S5

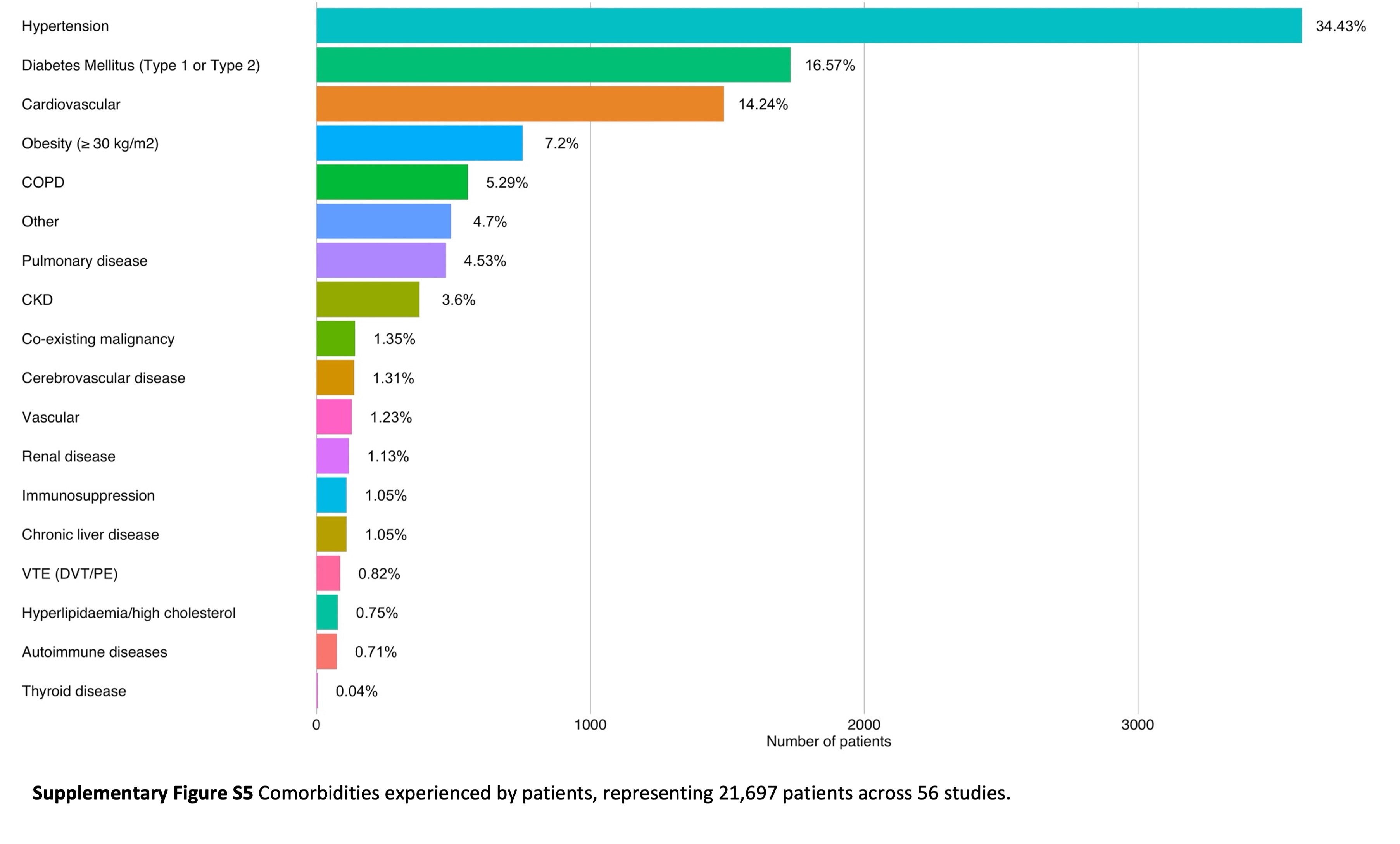

### Figure S6

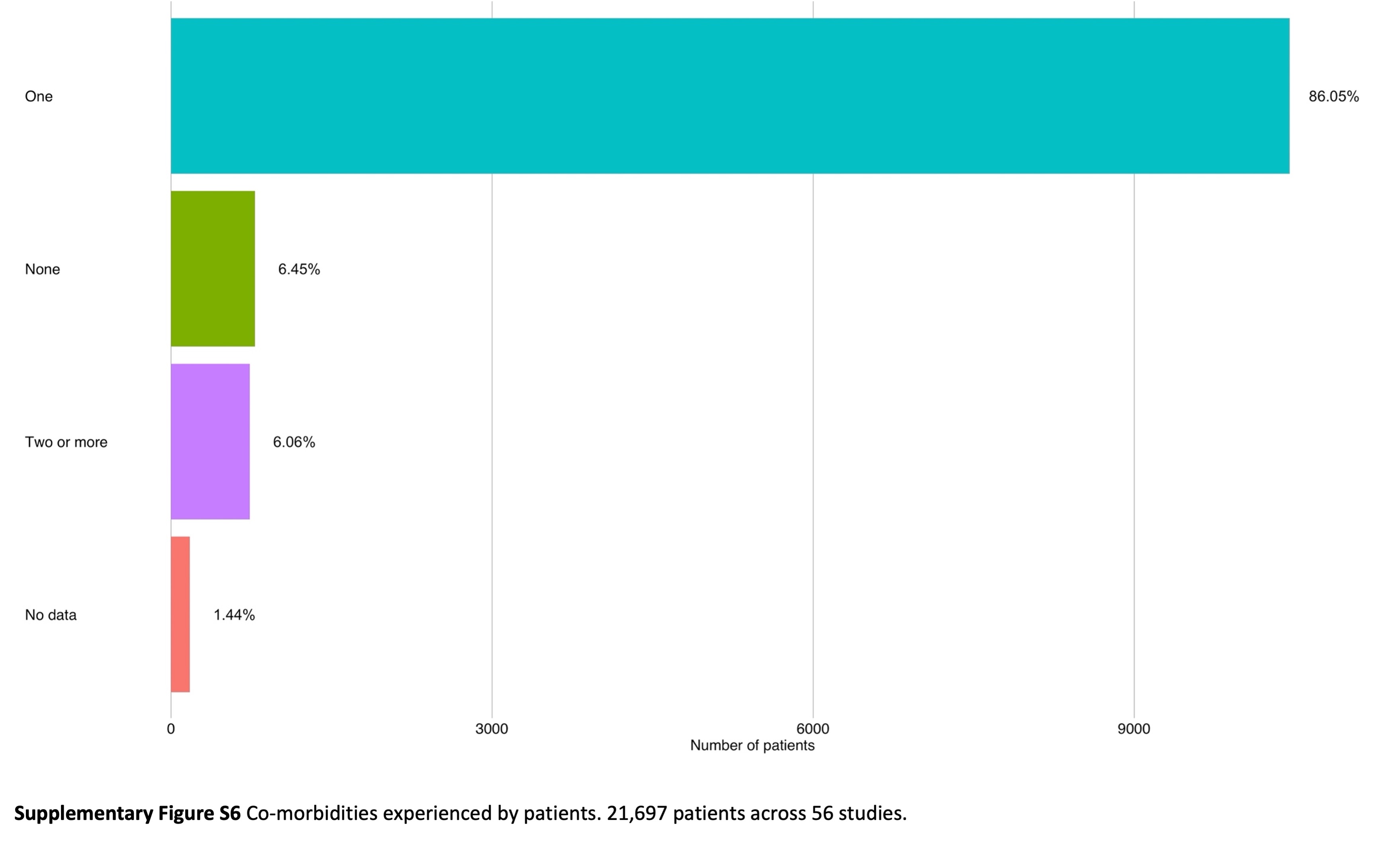

### Figure S7

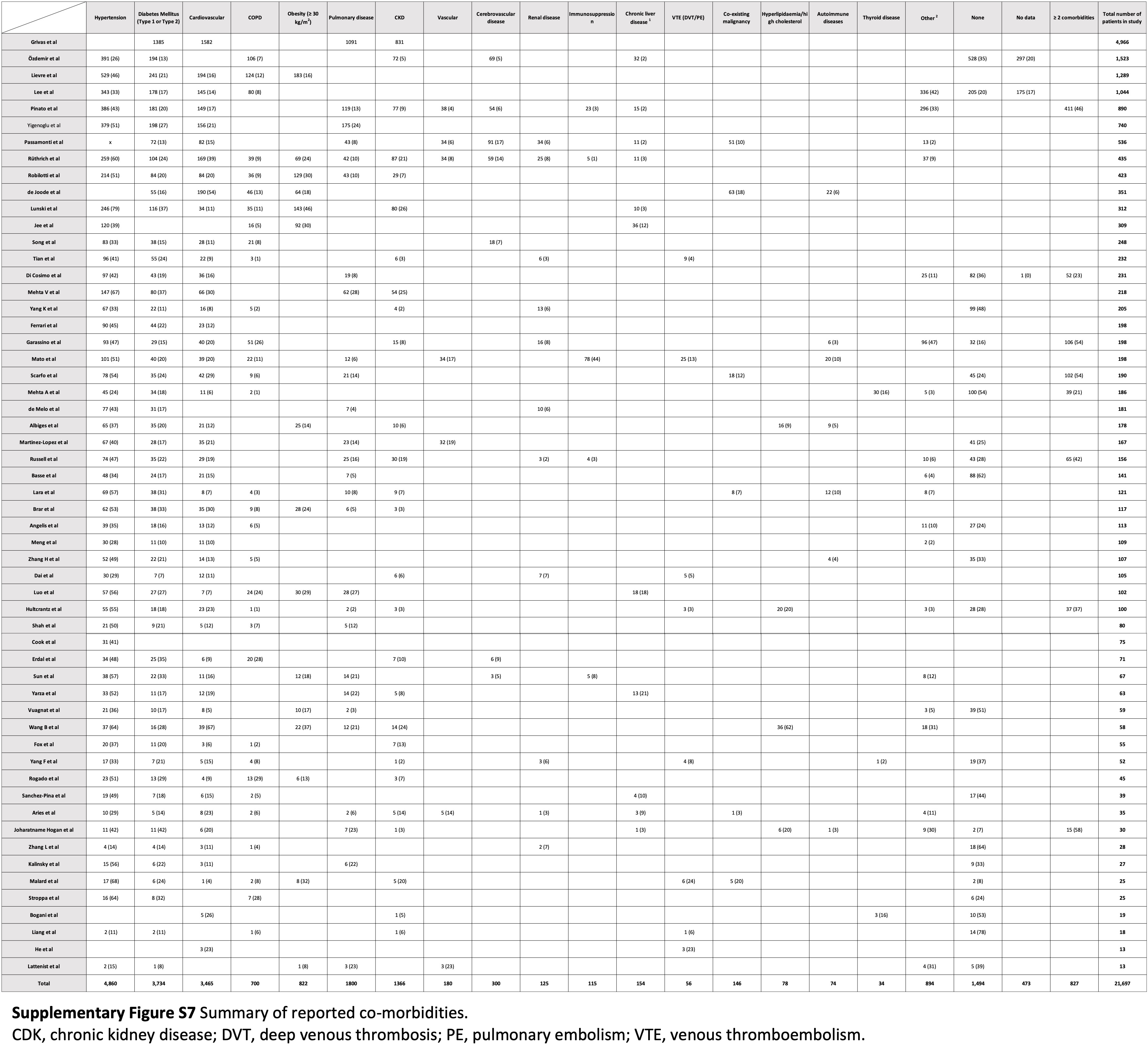

### Figure S8

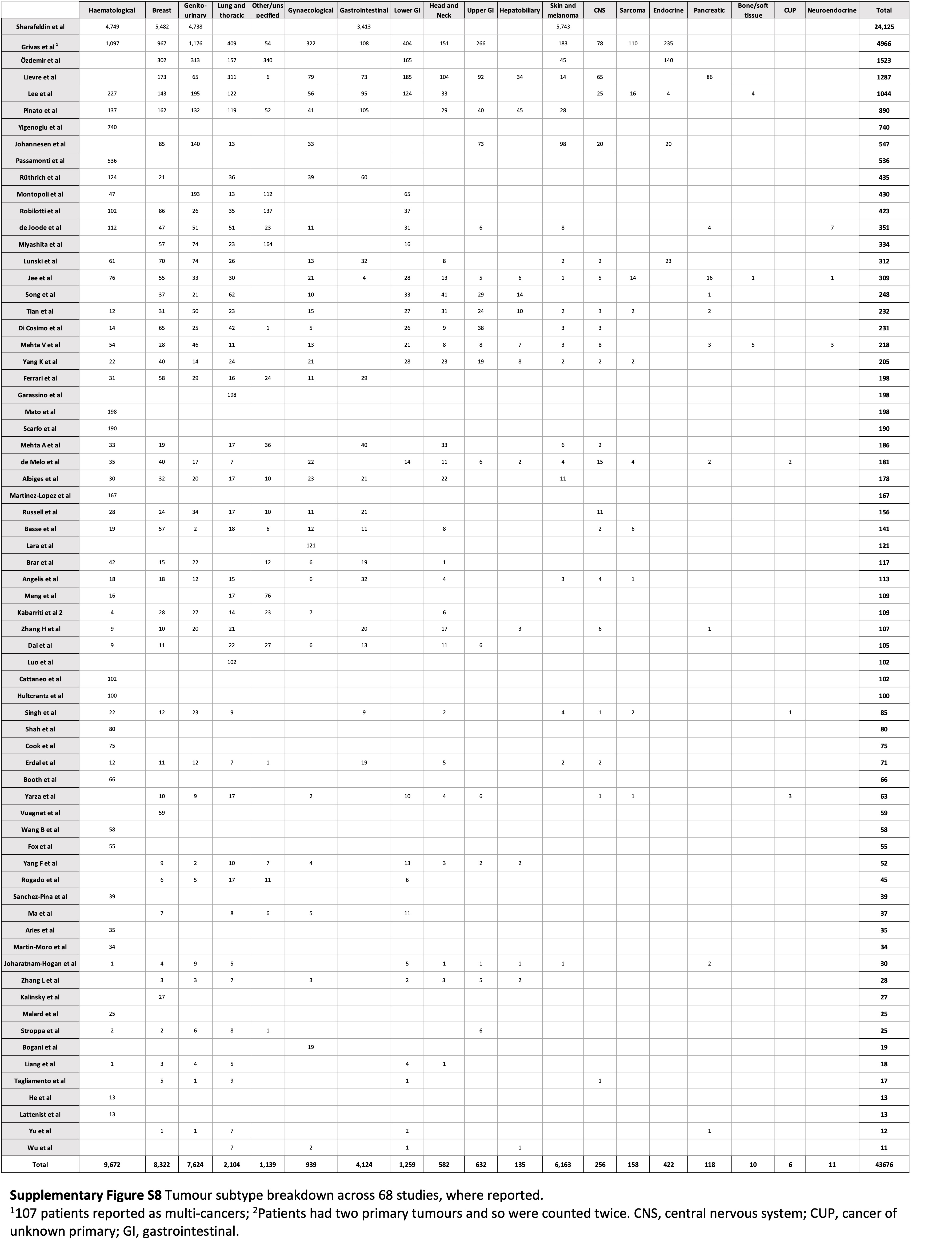

### Figure S9

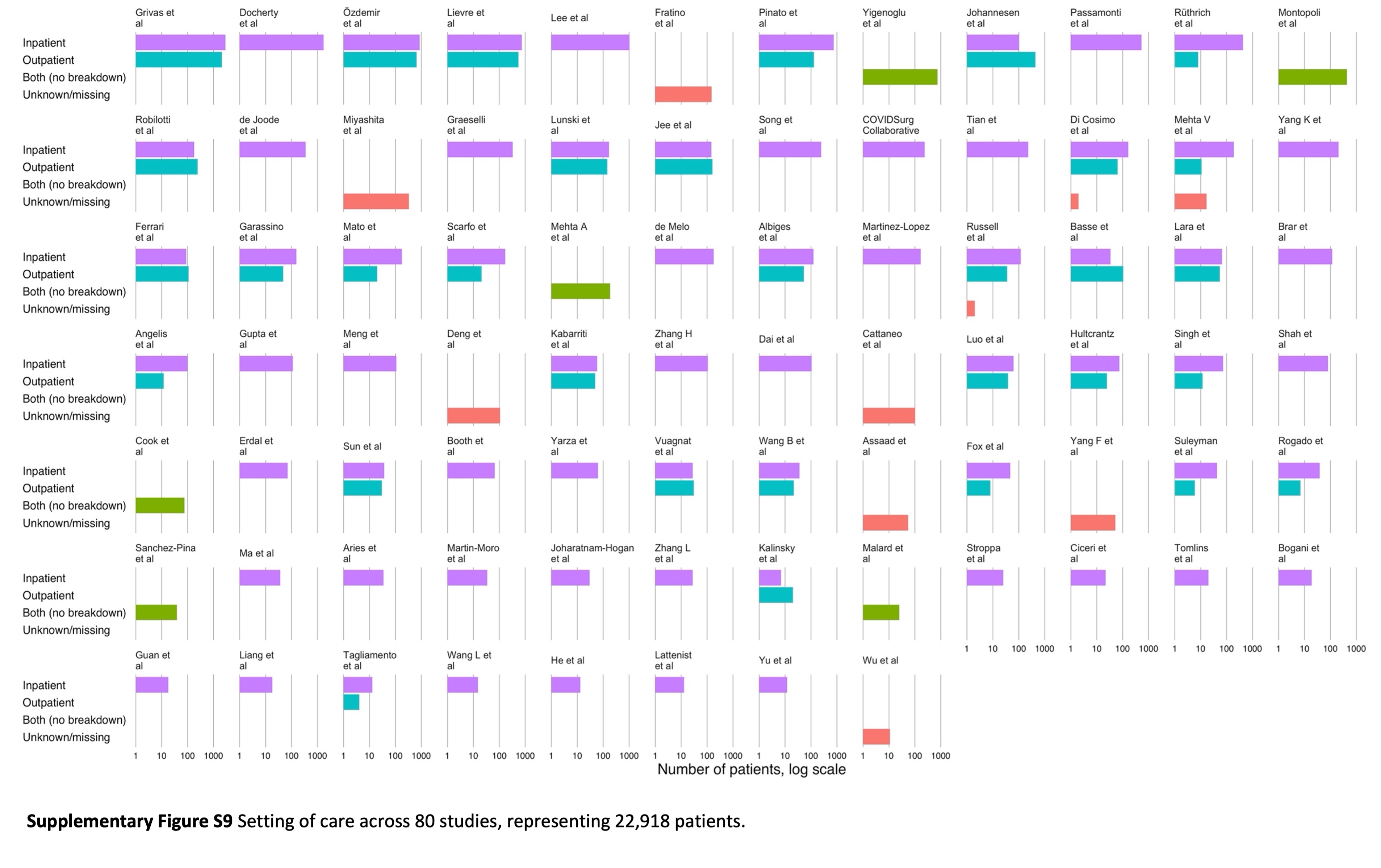

### Figure S10

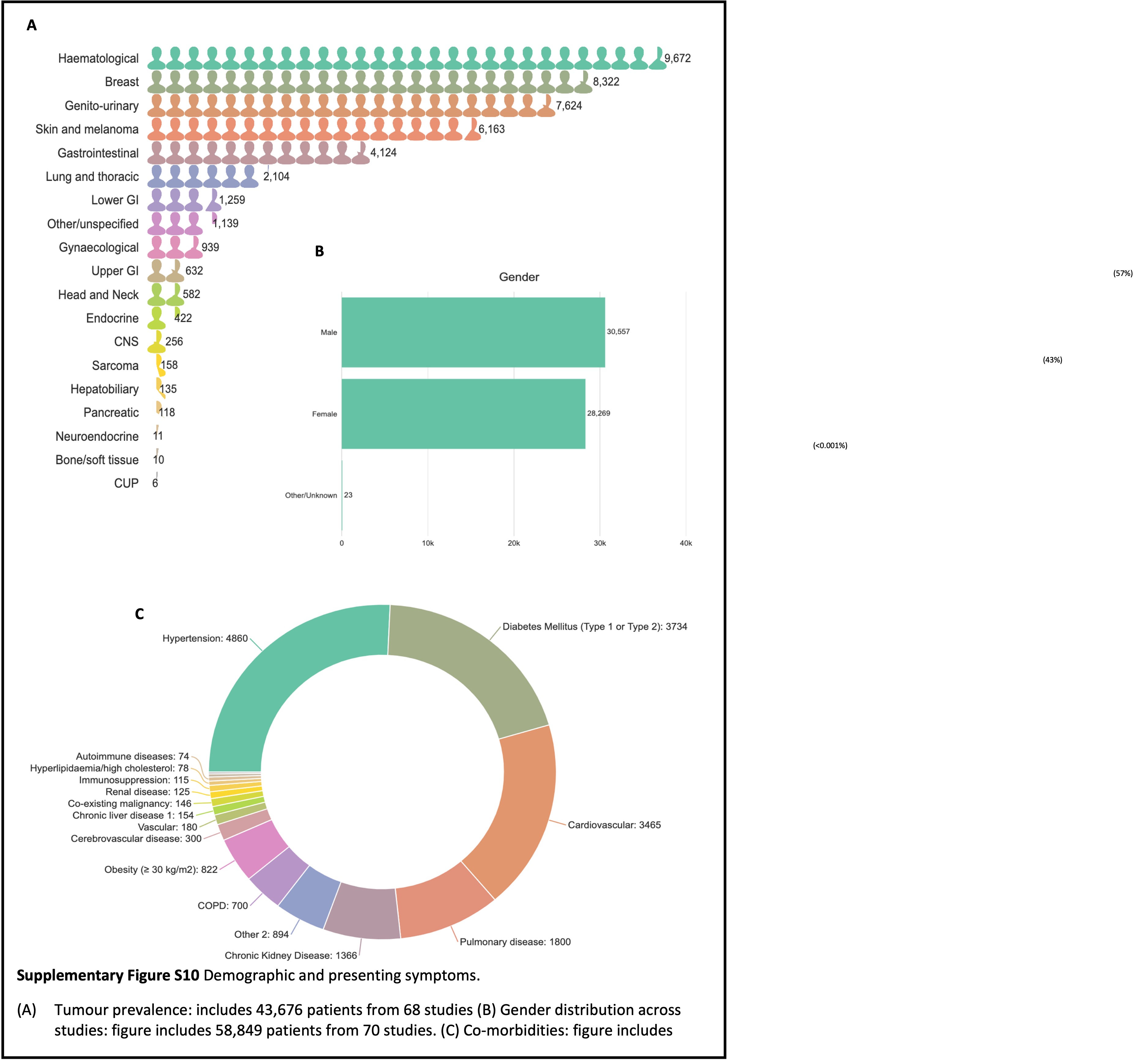

### Figure S11

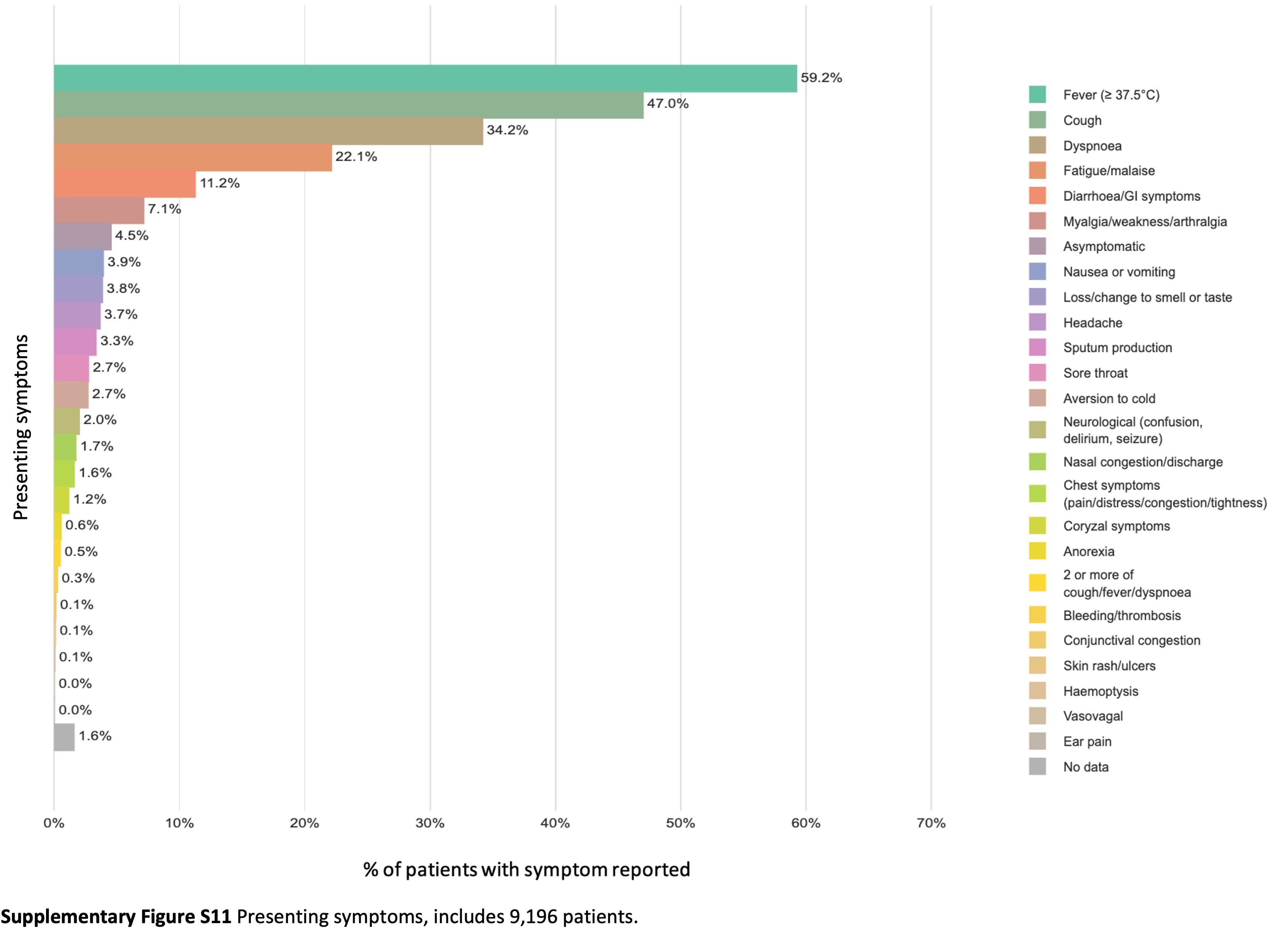

### Figure S12

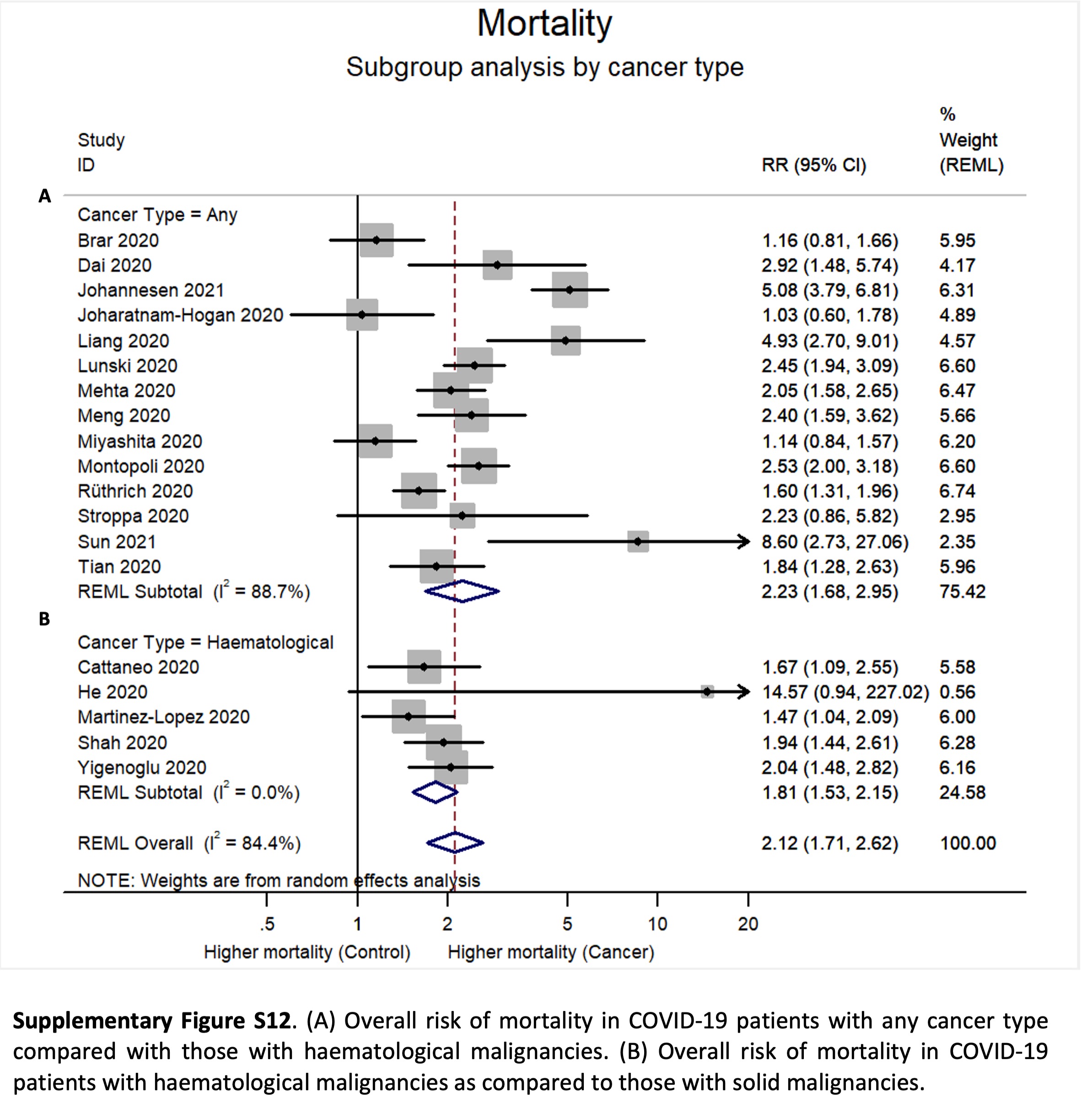

### Figure S13

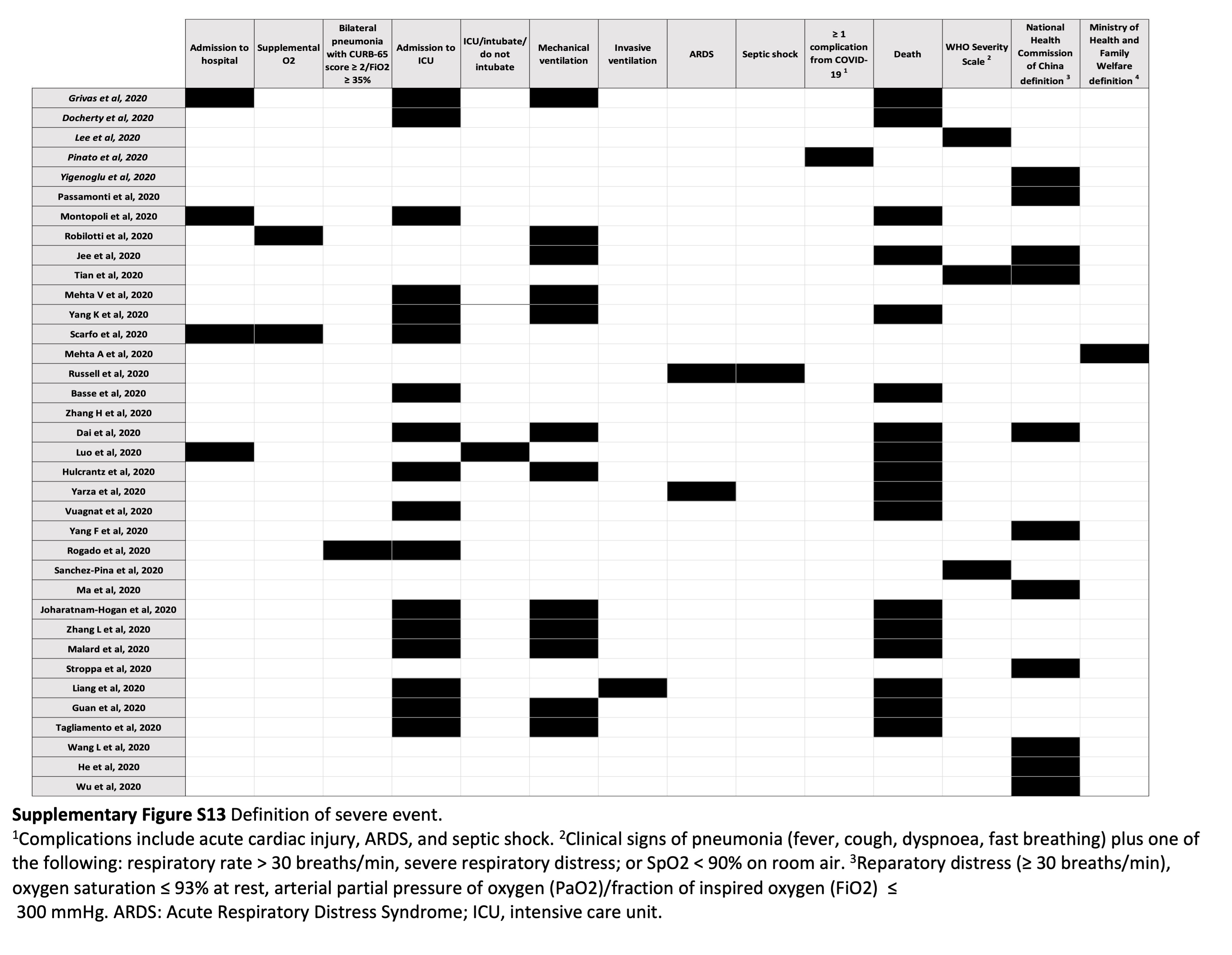

### Figure S14a

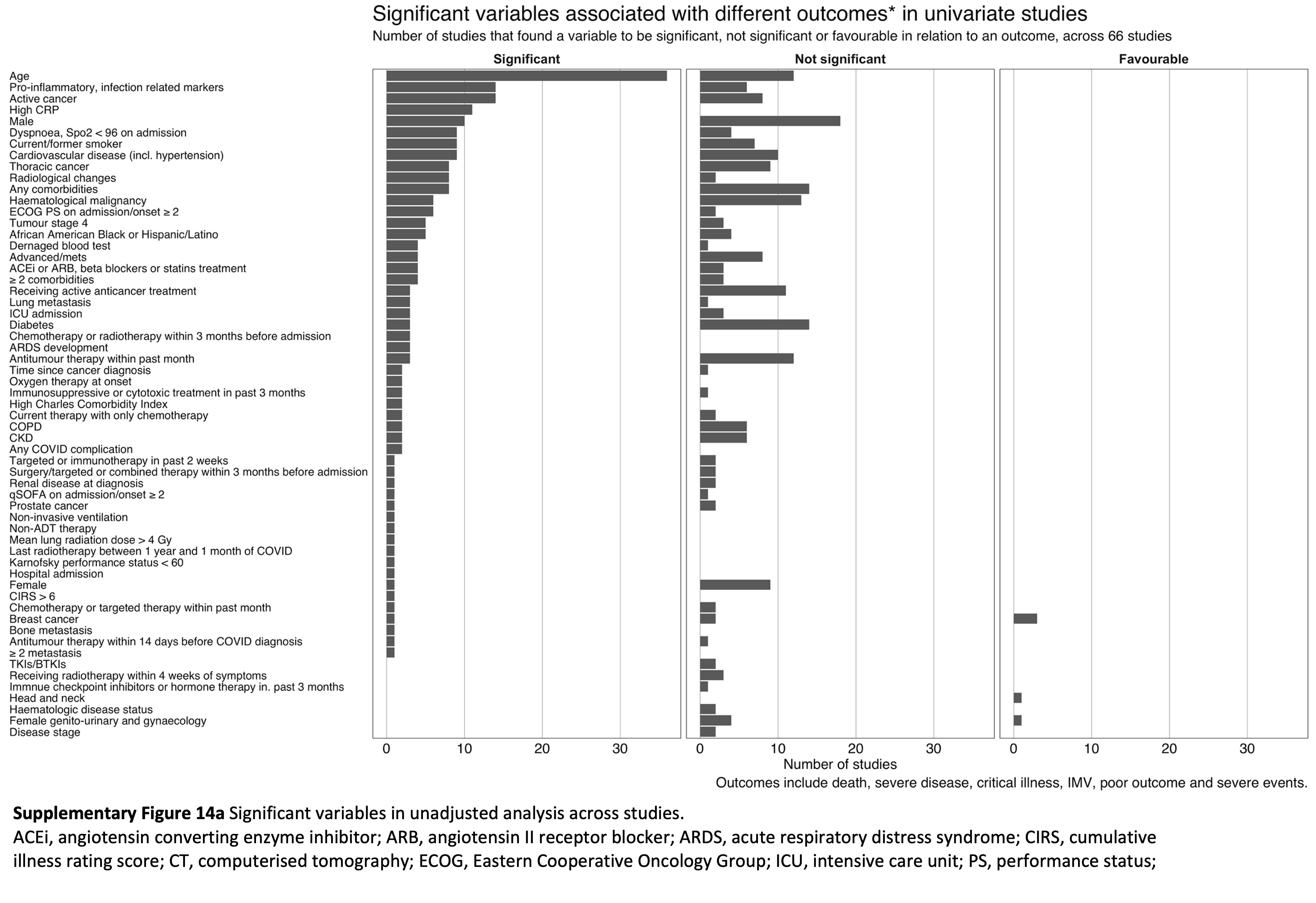

### Figure S14b

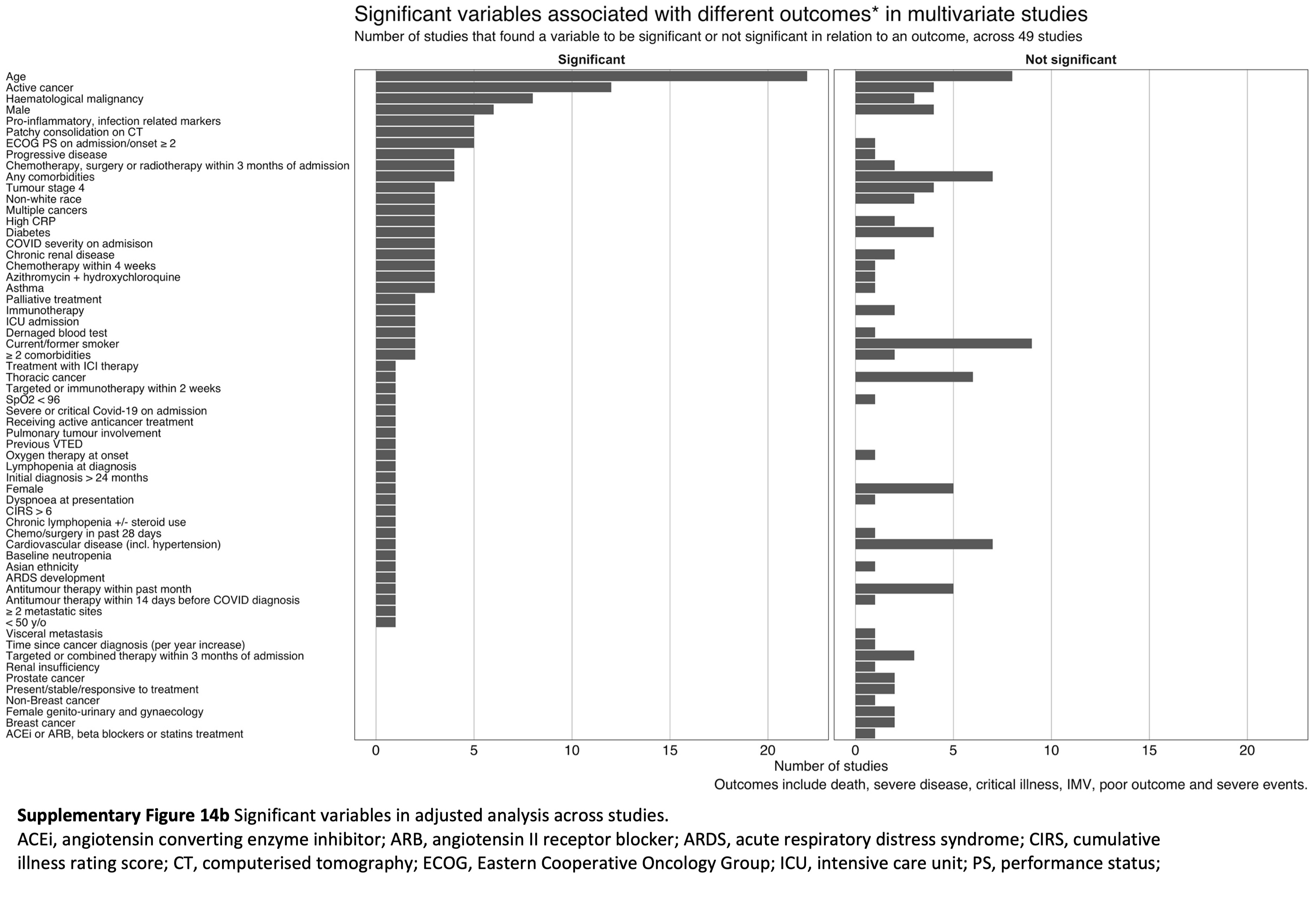

### Figure S15

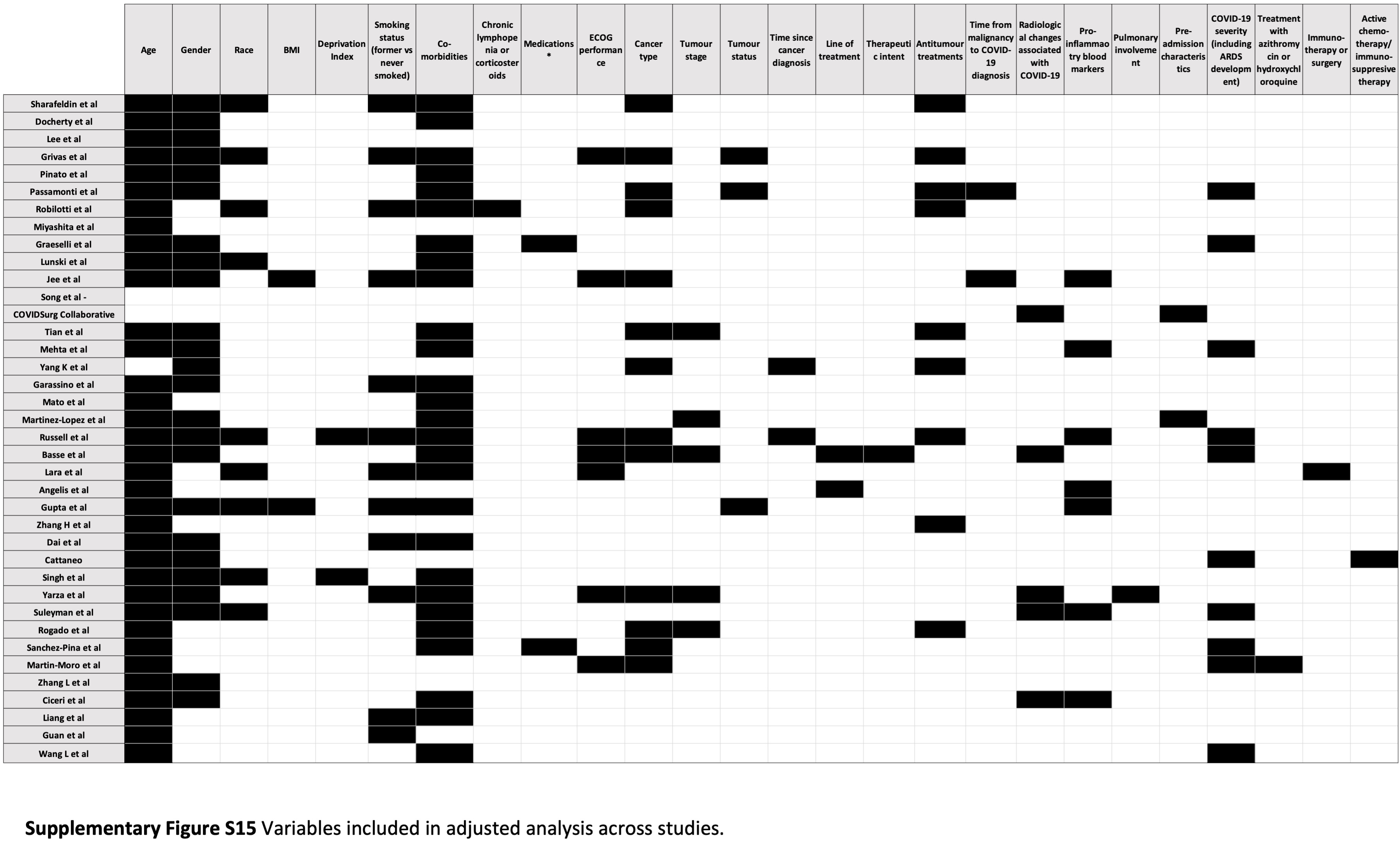

### Figure S16

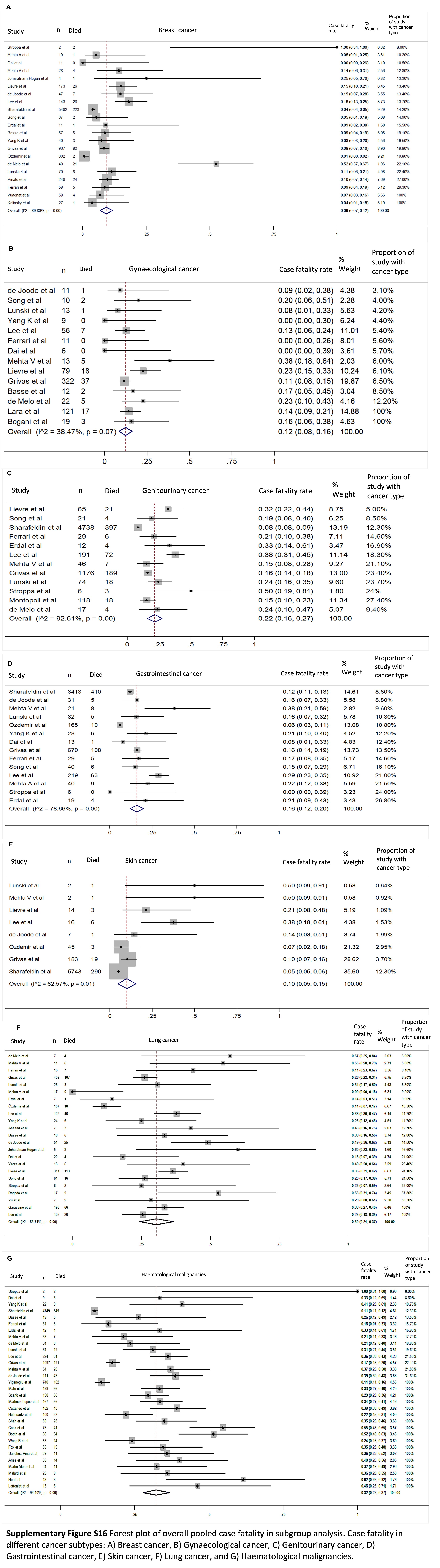

### Figure S17

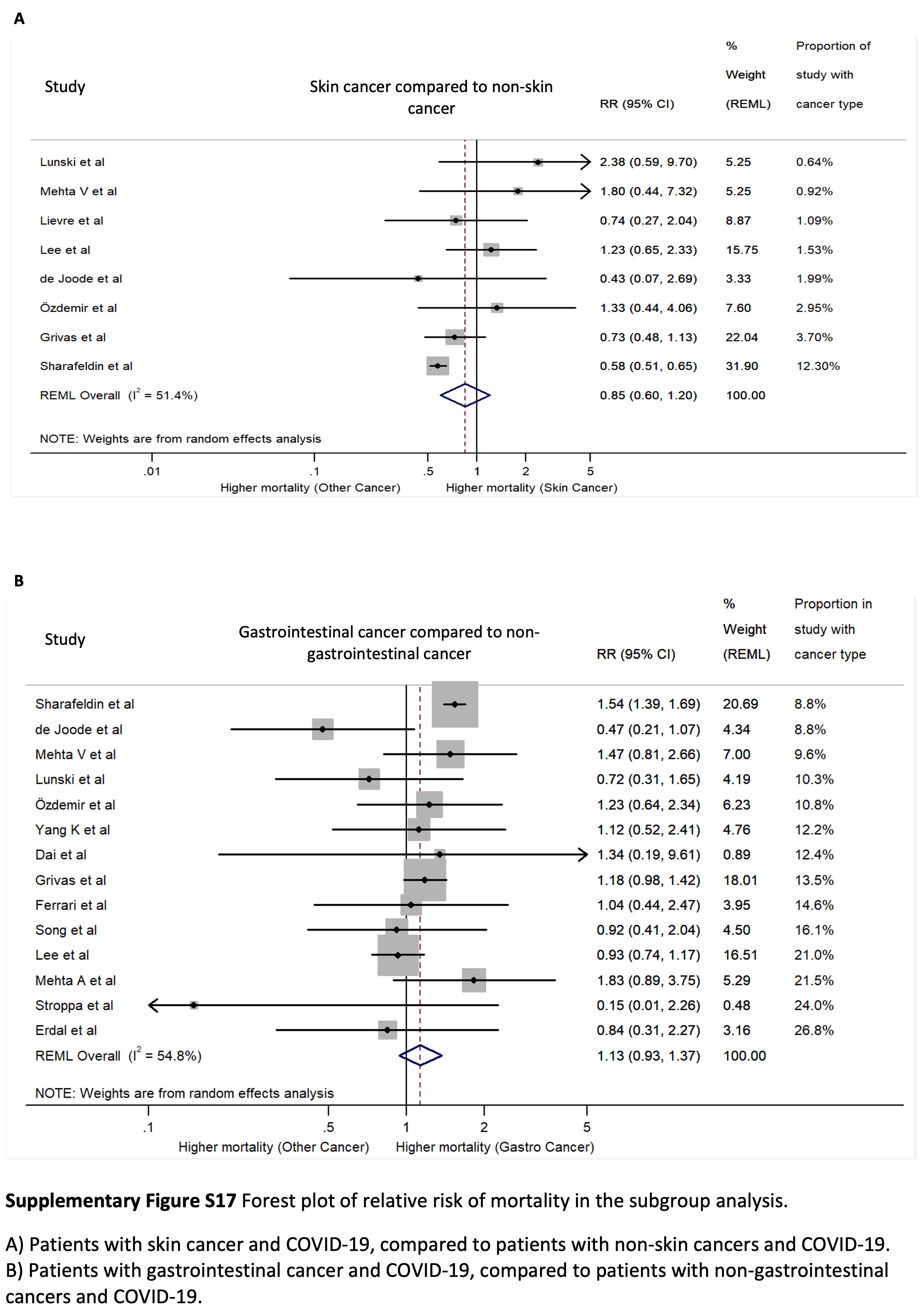

### Figure S18

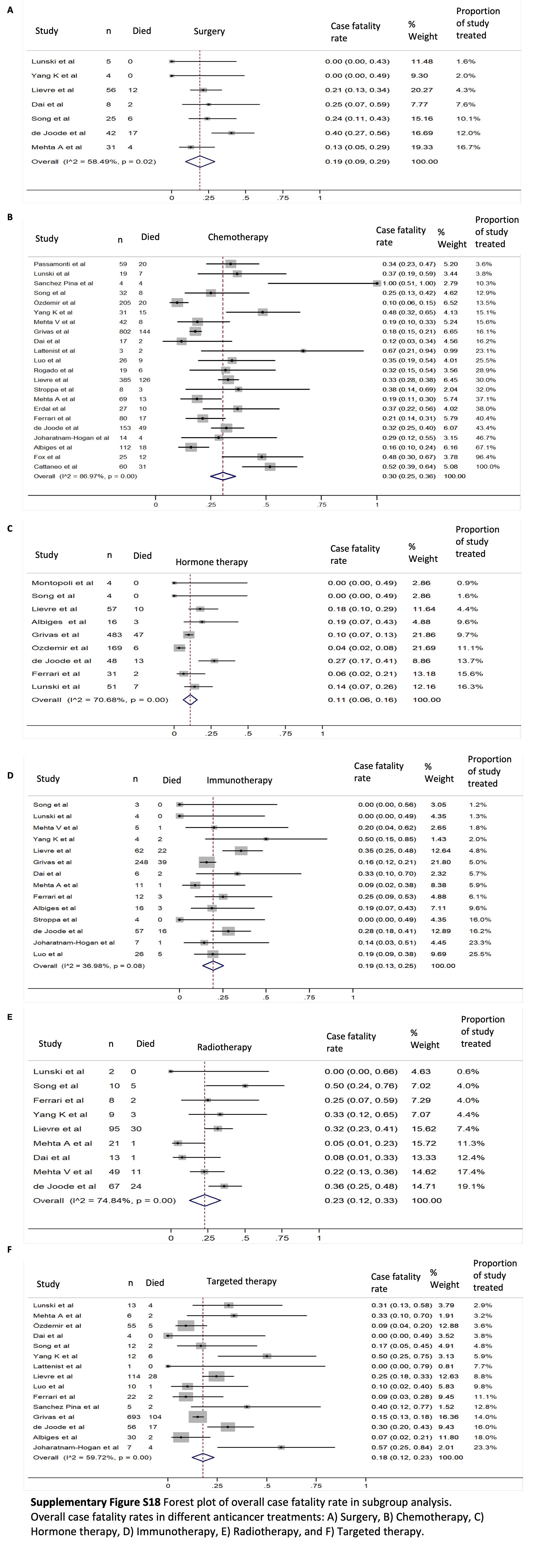
