## Supplementary material for "A comprehensive systematic review and meta-analysis of the global data involving 61,532 cancer patients with SARS-CoV-2 infection": Table S1

| **Search number** | **Query** | **Search Details** | **Results** | **Time** | **Date** |
| --- | --- | --- | --- | --- | --- |
| **14** | (((((cancer) OR (malignan*)) OR (tumour)) OR (neoplasm)) AND ((((2019-nCoV pneumonia) OR (SARS-CoV-2)) OR (Coronavirus)) OR (COVID-19))) AND ((solid) OR (haematolog*)) | ("cancer s"[All Fields] OR "cancerated"[All Fields] OR "canceration"[All Fields] OR "cancerization"[All Fields] OR "cancerized"[All Fields] OR "cancerous"[All Fields] OR "neoplasms"[MeSH Terms] OR "neoplasms"[All Fields] OR "cancer"[All Fields] OR "cancers"[All Fields] OR "malignan*"[All Fields] OR ("cysts"[MeSH Terms] OR "cysts"[All Fields] OR "cyst"[All Fields] OR "neurofibroma"[MeSH Terms] OR "neurofibroma"[All Fields] OR "neurofibromas"[All Fields] OR "tumor s"[All Fields] OR "tumoral"[All Fields] OR "tumorous"[All Fields] OR "tumour"[All Fields] OR "neoplasms"[MeSH Terms] OR "neoplasms"[All Fields] OR "tumor"[All Fields] OR "tumour s"[All Fields] OR "tumoural"[All Fields] OR "tumourous"[All Fields] OR "tumours"[All Fields] OR "tumors"[All Fields]) OR ("neoplasm s"[All Fields] OR "neoplasms"[MeSH Terms] OR "neoplasms"[All Fields] OR "neoplasm"[All Fields])) AND ((("severe acute respiratory syndrome coronavirus 2"[Supplementary Concept] OR "severe acute respiratory syndrome coronavirus 2"[All Fields] OR "2019 ncov"[All Fields]) AND ("pneumonia"[MeSH Terms] OR "pneumonia"[All Fields] OR "pneumoniae"[All Fields] OR "pneumonias"[All Fields] OR "pneumoniae s"[All Fields])) OR ("severe acute respiratory syndrome coronavirus 2"[Supplementary Concept] OR "severe acute respiratory syndrome coronavirus 2"[All Fields] OR "sars cov 2"[All Fields]) OR ("coronavirus"[MeSH Terms] OR "coronavirus"[All Fields] OR "coronaviruses"[All Fields]) OR ("severe acute respiratory syndrome coronavirus 2"[Supplementary Concept] OR "severe acute respiratory syndrome coronavirus 2"[All Fields] OR "ncov"[All Fields] OR "2019 ncov"[All Fields] OR "covid 19"[All Fields] OR "sars cov 2"[All Fields] OR (("coronavirus"[All Fields] OR "cov"[All Fields]) AND 2019/11/01:3000/12/31[Date - Publication]))) AND ("solid"[All Fields] OR "solid s"[All Fields] OR "solids"[All Fields] OR "haematolog*"[All Fields]) | 512 | 09:21:54 | 14/06/2021 |
| **13** | (solid) OR (haematolog*) | "solid"[All Fields] OR "solid s"[All Fields] OR "solids"[All Fields] OR "haematolog*"[All Fields] | 490,032 | 09:21:21 | 14/06/2021 |
| **12** | haematolog* | "haematolog*"[All Fields] | 90,344 | 09:20:30 | 14/06/2021 |
| **11** | solid | "solid"[All Fields] OR "solid s"[All Fields] OR "solids"[All Fields] | 402,191 | 09:20:00 | 14/06/2021 |
| **10** | (((2019-nCoV pneumonia) OR (SARS-CoV-2)) OR (Coronavirus)) OR (COVID-19) | (("severe acute respiratory syndrome coronavirus 2"[Supplementary Concept] OR "severe acute respiratory syndrome coronavirus 2"[All Fields] OR "2019 ncov"[All Fields]) AND ("pneumonia"[MeSH Terms] OR "pneumonia"[All Fields] OR "pneumoniae"[All Fields] OR "pneumonias"[All Fields] OR "pneumoniae s"[All Fields])) OR ("severe acute respiratory syndrome coronavirus 2"[Supplementary Concept] OR "severe acute respiratory syndrome coronavirus 2"[All Fields] OR "sars cov 2"[All Fields]) OR ("coronavirus"[MeSH Terms] OR "coronavirus"[All Fields] OR "coronaviruses"[All Fields]) OR ("severe acute respiratory syndrome coronavirus 2"[Supplementary Concept] OR "severe acute respiratory syndrome coronavirus 2"[All Fields] OR "ncov"[All Fields] OR "2019 ncov"[All Fields] OR "covid 19"[All Fields] OR "sars cov 2"[All Fields] OR (("coronavirus"[All Fields] OR "cov"[All Fields]) AND 2019/11/01:3000/12/31[Date - Publication])) | 161,008 | 09:19:24 | 14/06/2021 |
| **9** | COVID-19 | "severe acute respiratory syndrome coronavirus 2"[Supplementary Concept] OR "severe acute respiratory syndrome coronavirus 2"[All Fields] OR "ncov"[All Fields] OR "2019 ncov"[All Fields] OR "covid 19"[All Fields] OR "sars cov 2"[All Fields] OR (("coronavirus"[All Fields] OR "cov"[All Fields]) AND 2019/11/01:3000/12/31[Date - Publication]) | 145,752 | 09:19:08 | 14/06/2021 |
| **8** | coronavirus | "coronavirus"[MeSH Terms] OR "coronavirus"[All Fields] OR "coronaviruses"[All Fields] | 116,456 | 09:19:00 | 14/06/2021 |
| **7** | SARS-CoV-2 | "severe acute respiratory syndrome coronavirus 2"[Supplementary Concept] OR "severe acute respiratory syndrome coronavirus 2"[All Fields] OR "sars cov 2"[All Fields] | 89,189 | 09:18:53 | 14/06/2021 |
| **6** | 2019-nCoV pneumonia | ("severe acute respiratory syndrome coronavirus 2"[Supplementary Concept] OR "severe acute respiratory syndrome coronavirus 2"[All Fields] OR "2019 ncov"[All Fields]) AND ("pneumonia"[MeSH Terms] OR "pneumonia"[All Fields] OR "pneumoniae"[All Fields] OR "pneumonias"[All Fields] OR "pneumoniae s"[All Fields]) | 71,085 | 09:18:43 | 14/06/2021 |
| **5** | (((cancer) OR (malignan*)) OR (tumour)) OR (neoplasm) | "cancer s"[All Fields] OR "cancerated"[All Fields] OR "canceration"[All Fields] OR "cancerization"[All Fields] OR "cancerized"[All Fields] OR "cancerous"[All Fields] OR "neoplasms"[MeSH Terms] OR "neoplasms"[All Fields] OR "cancer"[All Fields] OR "cancers"[All Fields] OR "malignan*"[All Fields] OR "cysts"[MeSH Terms] OR "cysts"[All Fields] OR "cyst"[All Fields] OR "neurofibroma"[MeSH Terms] OR "neurofibroma"[All Fields] OR "neurofibromas"[All Fields] OR "tumor s"[All Fields] OR "tumoral"[All Fields] OR "tumorous"[All Fields] OR "tumour"[All Fields] OR "neoplasms"[MeSH Terms] OR "neoplasms"[All Fields] OR "tumor"[All Fields] OR "tumour s"[All Fields] OR "tumoural"[All Fields] OR "tumourous"[All Fields] OR "tumours"[All Fields] OR "tumors"[All Fields] OR "neoplasm s"[All Fields] OR "neoplasms"[MeSH Terms] OR "neoplasms"[All Fields] OR "neoplasm"[All Fields] | 4,997,027 | 09:18:33 | 14/06/2021 |
| **4** | neoplasm | "neoplasm s"[All Fields] OR "neoplasms"[MeSH Terms] OR "neoplasms"[All Fields] OR "neoplasm"[All Fields] | 3,558,546 | 09:18:19 | 14/06/2021 |
| **3** | tumour | "cysts"[MeSH Terms] OR "cysts"[All Fields] OR "cyst"[All Fields] OR "neurofibroma"[MeSH Terms] OR "neurofibroma"[All Fields] OR "neurofibromas"[All Fields] OR "tumor s"[All Fields] OR "tumoral"[All Fields] OR "tumorous"[All Fields] OR "tumour"[All Fields] OR "neoplasms"[MeSH Terms] OR "neoplasms"[All Fields] OR "tumor"[All Fields] OR "tumour s"[All Fields] OR "tumoural"[All Fields] OR "tumourous"[All Fields] OR "tumours"[All Fields] OR "tumors"[All Fields] | 4,274,667 | 09:18:14 | 14/06/2021 |
| **2** | malignan* | "malignan*"[All Fields] | 621,529 | 09:18:07 | 14/06/2021 |
| **1** | cancer | "cancer s"[All Fields] OR "cancerated"[All Fields] OR "canceration"[All Fields] OR "cancerization"[All Fields] OR "cancerized"[All Fields] OR "cancerous"[All Fields] OR "neoplasms"[MeSH Terms] OR "neoplasms"[All Fields] OR "cancer"[All Fields] OR "cancers"[All Fields] | 4,373,309 | 09:18:01 | 14/06/2021 |

**Supplementary Table S1** Search strategy used for the systematic review in PubMed.
