## Supplementary material for "A comprehensive systematic review and meta-analysis of the global data involving 61,532 cancer patients with SARS-CoV-2 infection": Table S2

| **Set** | **Results** | **Search History** | **Date** |
| --- | --- | --- | --- |
| #14 | 232 | #13 AND #10 AND #5  *Indexes=SCI-EXPANDED, SSCI, A&HCI, CPCI-S, CPCI-SSH, BKCI-S, BKCI-SSH, ESCI, CCR-EXPANDED, IC Timespan=All years* | 14/06/2021 |
| #13 | 1,402,806 | #12 OR #11  *Indexes=SCI-EXPANDED, SSCI, A&HCI, CPCI-S, CPCI-SSH, BKCI-S, BKCI-SSH, ESCI, CCR-EXPANDED, IC Timespan=All years* | 14/06/2021 |
| #12 | 29,734 | **TOPIC:** (haematolog*)  *Indexes=SCI-EXPANDED, SSCI, A&HCI, CPCI-S, CPCI-SSH, BKCI-S, BKCI-SSH, ESCI, CCR-EXPANDED, IC Timespan=All years* | 14/06/2021 |
| #11 | 1,374,645 | **TOPIC:** (solid)  *Indexes=SCI-EXPANDED, SSCI, A&HCI, CPCI-S, CPCI-SSH, BKCI-S, BKCI-SSH, ESCI, CCR-EXPANDED, IC Timespan=All years* | 14/06/2021 |
| #10 | 156,260 | #9 OR #8 OR #7 OR #6  *Indexes=SCI-EXPANDED, SSCI, A&HCI, CPCI-S, CPCI-SSH, BKCI-S, BKCI-SSH, ESCI, CCR-EXPANDED, IC Timespan=All years* | 14/06/2021 |
| #9 | 128,800 | **TOPIC:** (COVID-19)  *Indexes=SCI-EXPANDED, SSCI, A&HCI, CPCI-S, CPCI-SSH, BKCI-S, BKCI-SSH, ESCI, CCR-EXPANDED, IC Timespan=All years* | 14/06/2021 |
| #8 | 69,653 | **TOPIC:** (Coronavirus)  *Indexes=SCI-EXPANDED, SSCI, A&HCI, CPCI-S, CPCI-SSH, BKCI-S, BKCI-SSH, ESCI, CCR-EXPANDED, IC Timespan=All years* | 14/06/2021 |
| #7 | 39,067 | **TOPIC:** (SARS-CoV-2)  *Indexes=SCI-EXPANDED, SSCI, A&HCI, CPCI-S, CPCI-SSH, BKCI-S, BKCI-SSH, ESCI, CCR-EXPANDED, IC Timespan=All years* | 14/06/2021 |
| #6 | 498 | **TOPIC:** (2019-nCoV pneumonia)  *Indexes=SCI-EXPANDED, SSCI, A&HCI, CPCI-S, CPCI-SSH, BKCI-S, BKCI-SSH, ESCI, CCR-EXPANDED, IC Timespan=All years* | 14/06/2021 |
| #5 | 3,929,534 | #4 OR #3 OR #2 OR #1  *Indexes=SCI-EXPANDED, SSCI, A&HCI, CPCI-S, CPCI-SSH, BKCI-S, BKCI-SSH, ESCI, CCR-EXPANDED, IC Timespan=All years* | 14/06/2021 |
| #4 | 201,475 | **TOPIC:** (neoplasm)  *Indexes=SCI-EXPANDED, SSCI, A&HCI, CPCI-S, CPCI-SSH, BKCI-S, BKCI-SSH, ESCI, CCR-EXPANDED, IC Timespan=All years* | 14/06/2021 |
| #3 | 1,906,360 | T**OPIC:** (tumour)  *Indexes=SCI-EXPANDED, SSCI, A&HCI, CPCI-S, CPCI-SSH, BKCI-S, BKCI-SSH, ESCI, CCR-EXPANDED, IC Timespan=All years* | 14/06/2021 |
| #2 | 604,111 | **TOPIC:** (malignan*)  *Indexes=SCI-EXPANDED, SSCI, A&HCI, CPCI-S, CPCI-SSH, BKCI-S, BKCI-SSH, ESCI, CCR-EXPANDED, IC Timespan=All years* | 14/06/2021 |
| # 1 | 2,709,682 | **TOPIC:** (cancer)  ***Indexes=SCI-EXPANDED, SSCI, A&HCI, CPCI-S, CPCI-SSH, BKCI-S, BKCI-SSH, ESCI, CCR-EXPANDED, IC Timespan=All years*** | 14/06/2021 |

**Supplementary Table S2** Search strategy used for the systematic review in Web of Science.
