## Supplementary material for "A comprehensive systematic review and meta-analysis of the global data involving 61,532 cancer patients with SARS-CoV-2 infection": Table S3

| **Search Number** | **Search History** | **Document results** | **Date** |
| --- | --- | --- | --- |
| **14** | ( ( TITLE-ABS-KEY ( cancer ) )  OR  ( TITLE-ABS-KEY ( malignan* ) )  OR  ( TITLE-ABS-KEY ( tumour ) )  OR  ( TITLE-ABS-KEY ( neoplasm ) ) )  AND  ( ( 2019-ncov  AND pneumonia )  OR  ( sars-cov-2 )  OR  ( coronavirus )  OR  ( covid-19 ) )  AND  ( ( solid )  OR  ( haematolog* ) ) | 364 | 14/06/2021 |
| **13** | ( solid )  OR  ( haematolog* ) | 1,797,587 | 14/06/2021 |
| **12** | haematolog* | 44,098 | 14/06/2021 |
| **11** | solid | 1,755,385 | 14/06/2021 |
| **10** | ( 2019-ncov  AND pneumonia )  OR  ( sars-cov-2 )  OR  ( coronavirus )  OR  ( covid-19 ) | 192,773 | 14/06/2021 |
| **9** | covid-19 | 155,108 | 14/06/2021 |
| **8** | coronavirus | 134,571 | 14/06/2021 |
| **7** | sars-cov-2 | 63,082 | 14/06/2021 |
| **6** | 2019-ncov AND pneumonia | 988 | 14/06/2021 |
| **5** | ( TITLE-ABS-KEY ( cancer ) )  OR  ( TITLE-ABS-KEY ( malignan* ) )  OR  ( TITLE-ABS-KEY ( tumour ) )  OR  ( TITLE-ABS-KEY ( neoplasm ) ) | 5,338,087 | 14/06/2021 |
| **4** | neoplasm | 2,712,836 | 14/06/2021 |
| **3** | tumour | 3,447,845 | 14/06/2021 |
| **2** | malignan* | 805,290 | 14/06/2021 |
| **1** | Cancer | 3,358,220 | 14/06/2021 |

**Supplementary Table S3** Search strategy used for the systematic review in Scopus.
