## Supplementary material for "A comprehensive systematic review and meta-analysis of the global data involving 61,532 cancer patients with SARS-CoV-2 infection": Table S4

| **Publication** | **Country** | **Study type** | **Time period** | **Cancer patients** | **Cancer type** | **Cancer vs non cancer cohort** | **Presenting symptoms** | **Co- morbidities** | **Impact of cancer treatment** | **Severity** | **Outcome** |
| --- | --- | --- | --- | --- | --- | --- | --- | --- | --- | --- | --- |
| Sharafeldin et al | USA | N3C Cohort, multicentre, retrospective | 1st January 2020 to 25th March 2021 | 38,614 | Any |  |  |  |  |  |  |
| Grivas et al * | Multicentre^1^ | Covid-19 and Cancer Consortium registry database | 17th March to 16th April 2020 | 4966 | Any |  |  |  |  |  |  |
| Docherty et al | United Kingdom | Multi-centre, prospective, 208 centres | 6th February to 19th April 2020 | 1743 | Any |  |  |  |  |  |  |
| Özdemir et al | Turkey | Multi centre, retro-prospective | 11th March to 20th May 2020 | 1523 | Solid cancers |  |  |  |  |  |  |
| Lièvre et al^2^ | France | Multi-centre, retro-prospective | 4^th^ April to 11^th^ June 2020 | 1289 | Solid cancers |  |  |  |  |  |  |
| Lee et al * | United Kingdom | Multi-centre, prospective | 18th March to 8th May 2020 | 1044 | Any |  |  |  |  |  |  |
| Fratino et al | Italy | Multi centre, retrospective | Up to 30th March 2020 | 909 | Any |  |  |  |  |  |  |
| Pinato et al | Multicentre^2^ | Multi-centre, 19 centres, retrospective | 26th February to 1st April 2020 | 890 | Any |  |  |  |  |  |  |
| Yigenoglu et al | Turkey | Multi centre, retrospective | 11th March to 22nd June 2020 | 740 | Haematological malignancies |  |  |  |  |  |  |
| Johannesen et al | Norway | Multi-centre, retrospective | 1st January to 31st May 2020 | 547 | Any |  |  |  |  |  |  |
| Passamonti et al | Italy | Multi-centre, retrospective | 25th February to 18th May 2020 | 536 | Haematological malignancies |  |  |  |  |  |  |
| Rüthrich et al | Germany | Multi centre, retrospective | 16th March to 31st August 2020 | 435 | Any |  |  |  |  |  |  |
| Montopoli et al | Italy | Multi-centre, retrospective, 68 centres | Up to 1st April 2020 | 430 | Any (male patients only) |  |  |  |  |  |  |
| Robilotti et al | USA | Single centre, retrospective | 10th March to 7th May 2020 | 423 | Any |  |  |  |  |  |  |
| de Joode et al ^9^ | The Netherlands | Multi-centre, retrospective | 27^th^ March to 4^th^ May 2020 | 351 | Any |  |  |  |  |  |  |
| Miyashita et al | USA | Single centre | 1st March to 6th April 2020 | 334 | Any |  |  |  |  |  |  |
| Graeselli et al | Italy | Multi-centre, retrospective | 20th February to 22nd April 2020 | 331 | Any |  |  |  |  |  |  |
| Lunski et al | USA | Multi centre, retrospective | 1st March to 30th April 2020 | 312 | Any |  |  |  |  |  |  |
| Jee et al | USA | Single centre, retrospective | 8th March to 31st March 2020 | 309 | Any |  |  |  |  |  |  |
| Song et al | China | Multi centre, 33 centres, retrospective | 1st January to 25th March 2020 | 248 | Any |  |  |  |  |  |  |
| COVIDSurg Collaborative | 24 countries^3^ | Multi centre | 1st January to 31st March 2020 | 239 | Any |  |  |  |  |  |  |
| Tian et al | China | Multi-centre, retrospective | 13th January to 18th March 2020 | 232 | Any |  |  |  |  |  |  |
| Di Cosimo et al | Italy | Multi centre, 26 centres, retro-prospective | 15th May to 30th September 2020 | 231 | Any |  |  |  |  |  |  |
| Mehta V et al | USA | Single centre, retrospective | 18th March to 8th April 2020 | 218 | Any |  |  |  |  |  |  |
| Yang K et al | China | Multi-centre, retrospective | 13th January to 18th March 2020 | 205 | Any |  |  |  |  |  |  |
| Ferrari et al | Brazil | Multi centre, prospective | 29th March to 4th July 2020 | 198 | Any |  |  |  |  |  |  |
| Garassino et al | Multicentre^4^ | Multi-centre longitudinal | 26th March to 12th April 2020 | 198 | Thoracic cancer |  |  |  |  |  |  |
| Mato et al | Multicentre^5^ | Multi-centre, 43 centres, retrospective | 17th February to 30th April 2020 | 198 | CLL |  |  |  |  |  |  |
| Scarfo et al | International centres^6^ | Multi-centre, retrospective | 28th March to 22nd May 2020 | 190 | CLL |  |  |  |  |  |  |
| Mehta A et al | India | Single centre, retrospective | 8th June to 20th August 2020 | 186 | Any |  |  |  |  |  |  |
| de Melo et al | Brazil | Single centre | 30th April to 26th May 2020 | 181 | Any |  |  |  |  |  |  |
| Albiges et al | France | Single centre | 24th March to 29th April 2020 | 178 | Any |  |  |  |  |  |  |
| Martinez-Lopez et al | Spain | Multi-centre, 73 centres, retrospective | 1st March to 30th April 2020 | 167 | Myeloma |  |  |  |  |  |  |
| Russell et al | United Kingdom | Single centre, retrospective | 29th February to 12th May 2020 | 156 | Any |  |  |  |  |  |  |
| Basse et al | France | Single centre, prospective | 13th March to 25th April 2020 | 141 | Any |  |  |  |  |  |  |
| Lara et al | USA | Multi centre, retrospective | 1st March to 22nd April 2020 | 121 | Gynaecological cancer |  |  |  |  |  |  |
| Brar et al ^29^ | USA | Single centre, retrospective | 3^rd^ March to 15^th^ May 2020 | 117 | Any |  |  |  |  |  |  |
| Angelis et al | United Kingdom | Single centre, prospective | 1st March to 30th April 2020 | 113 | Any |  |  |  |  |  |  |
| Gupta et al | USA | Multi centre, 65 ICU centres | 4th March to 4th April 2002 | 112 | Any |  |  |  |  |  |  |
| Meng et al | China | Single centre, retrospective | 18th January to 27th March 2020 | 109 | Any |  |  |  |  |  |  |
| Deng et al | China | Multi-centre, retrospective | Up to 11th February 2020 | 107 | Any |  |  |  |  |  |  |
| Kabarriti et al | USA | Single centre | 14th March to 15th April 2020 | 107 | Any (all prior radiotherapy to lung) |  |  |  |  |  |  |
| Zhang H et al | China | Multi centre, 5 centres, retrospective | 5th January to 18th March 2020 | 107 | Any |  |  |  |  |  |  |
| Dai et al | China | Multi-centre, retrospective | 1st January to 24th February 2020 | 105 | Any |  |  |  |  |  |  |
| Luo et al | USA | Single centre, retrospective | 12th March to 6th May 2020 | 102 | Lung cancer |  |  |  |  |  |  |
| Cattaneo et al | Italy | Multi centre, retrospective | 1st March to 31st March 2020 | 102 | Haematological malignancies |  |  |  |  |  |  |
| Hultcrantz et al | USA | Multi-centre, 5 centres, retrospective | 1st March to 30th April 2020 | 100 | Multiple myeloma |  |  |  |  |  |  |
| Singh et al | USA | Single centre, retrospective | 10th March to 17th April 2020 | 85 | Any |  |  |  |  |  |  |
| Shah et al ^41^ | United Kingdom | Multi-centre, retrospective | 13^th^ March to 5^th^ May 2020 | 80 | Haematological malignancies |  |  |  |  |  |  |
| Cook et al | United Kingdom | Multicentre, prospective | Up to 18th May 2020 | 75 | Multiple myeloma |  |  |  |  |  |  |
| Erdal et al ^43^ | Turkey | Single centre, retrospective | 15^th^ March to 15^th^ May 2020 | 71 | Any |  |  |  |  |  |  |
| Sun et al ^44^ | USA | Multi-centre, retrospective | Up to June 2020 | 67 | Any |  |  |  |  |  |  |
| Booth et al | United Kingdom | Multi centre, prospective | 1st March to 6th May 2020 | 66 | Haematological malignancies |  |  |  |  |  |  |
| Yarza et al | Spain | Single centre, prospective | 9th March to 19th April 2020 | 63 | Any |  |  |  |  |  |  |
| Vuagnat et al | France | Single centre, prospective | 13th March to 25th April 2020 | 59 | Breast cancer |  |  |  |  |  |  |
| Wang B et al | USA | Single centre, retrospective | 1st March to 30th April 2020 | 58 | Multiple myeloma |  |  |  |  |  |  |
| Assaad et al | France | Multi-centre, prospective | 1st March to 25th April 2020 | 55 | Any |  |  |  |  |  |  |
| Fox et al | United Kingdom | Single centre, retrospective | 20th March to 20th April 2020 | 55 | Haematological malignancies |  |  |  |  |  |  |
| Yang F et al | China | Retrospective study | 1st January to 15th April 2020 | 52 | Any |  |  |  |  |  |  |
| Suleyman et al | USA | Multi centre, retrospective | 9th March t0 27th March 2020 | 49 | Any |  |  |  |  |  |  |
| Rogado et al | Spain | Single centre, retrospective | 1st February to 7th April 2020 | 45 | Any |  |  |  |  |  |  |
| Sanchez-Pina et al | Spain | Single centre, retrospective | 7th March to 7th April 2020 | 39 | Haematological malignancies |  |  |  |  |  |  |
| Ma et al | China | Single centre, retrospective | 1st January to 30th March 2020 | 37 | Any |  |  |  |  |  |  |
| Aries et al | United Kingdom | Single centre | 11th March to 11th May 2020 | 35 | Haematological malignancies |  |  |  |  |  |  |
| Martin-Moro et al | Spain | Single centre, retrospective | 9th March to 17th April 2020 | 34 | Haematological malignancies |  |  |  |  |  |  |
| Joharatnam-Hogan et al | United Kingdom | Multi centre, retrospective, 4 centres | 12th March to 7th April 2020 | 30 | Any |  |  |  |  |  |  |
| Zhang L et al | China | Multi-centre, retrospective | 13th January to 26th February 2020 | 28 | Any |  |  |  |  |  |  |
| Kalinsky et al | USA | Single centre | 10th March to 29th April 2020 | 27 | Breast cancer |  |  |  |  |  |  |
| Malard et al | France | Single centre | 9th March to 4th April 2020 | 25 | Haematological malignancies |  |  |  |  |  |  |
| Stroppa et al | Italy | Single centre | 21st February to 18th March 2020 | 25 | Any |  |  |  |  |  |  |
| Ciceri et al | Italy | Single centre | 25th February to 24th March 2020 | 22 | Any |  |  |  |  |  |  |
| Tomlins et al | United Kingdom | Single centre, retrospective | 10th March to 30th March 2020 | 20 | Any |  |  |  |  |  |  |
| Bogani et al | Italy | Single centre, retrospective | February to March 2020 | 19 | Gynaecological cancer |  |  |  |  |  |  |
| Liang et al | China | Multi-centre, prospective | 31st January 2020 | 18 | Any |  |  |  |  |  |  |
| Guan et al | China | Multi centre, retrospective, 575 hospitals | 11th December to 31st January 2020 | 18 | Any |  |  |  |  |  |  |
| Tagliamento et al | Italy | Single centre | 10th March to 6th April 2020 | 17 | Any |  |  |  |  |  |  |
| Wang L et al | China | Single centre, retrospective | 1st January to 6th February 2020 | 15 | Any |  |  |  |  |  |  |
| He et al | China | Multi centre | 23rd January to 14th February 2020 | 13 | Haematological malignancies |  |  |  |  |  |  |
| Lattenist et al | Belgium | Single centre, retrospective | 13th March to 15th May 2020 | 13 | Haematological malignancies |  |  |  |  |  |  |
| Yu et al | China | Single centre, retrospective | 30th December to 17th February 2020 | 12 | Any |  |  |  |  |  |  |
| Wu et al | China | Multi centre, retrospective, 2 centres | 9th January to 20th March 2020 | 11 | Any |  |  |  |  |  |  |

**Supplementary Table S4** Overview of studies included in review.

CCC-19, COVID-19 and Cancer Consortium; CLL, chronic lymphocytic leukaemia. Largest and most up to date study included in review. Shaded box indicates inclusion of factor.
