## Supplementary material for "A comprehensive systematic review and meta-analysis of the global data involving 61,532 cancer patients with SARS-CoV-2 infection": Table S5

| **Country** | **Number of patients** |
| --- | --- |
| **USA** | 41,423 |
| **United Kingdom** | 3,833 |
| **Italy** | 2,495 |
| **France** | 1,793 |
| **Turkey** | 2.345 |
| **China** | 1,318 |
| **Spain** | 934 |
| **Norway** | 547 |
| **Germany** | 447 |
| **Brazil** | 379 |
| **The Netherlands** | 363 |
| **India** | 186 |
| **Belgium** | 21 |
| **Ireland** | 18 |
| **Portugal** | 16 |
| **Switzerland** | 15 |
| **Egypt** | 12 |
| **Greece** | 8 |
| **Libya** | 4 |
| **Croatia** | 3 |
| **Algeria** | 11 |
| **Azerbaijan** | 1 |
| **Denmark** | 1 |
| **Israel** | 1 |
| **Jordan** | 1 |
| **Mexico** | 1 |
| **Pakistan** | 1 |
| **Sudan** | 1 |
| **Total** | **56,168** |

**Supplementary Table S5** Distribution of patients.

56,168 patients included across 80 studies.
