## Supplementary material for "A comprehensive systematic review and meta-analysis of the global data involving 61,532 cancer patients with SARS-CoV-2 infection": Table S6

| **Cancer subtype** | **Number of patients** | **% of total** |
| --- | --- | --- |
| **Haematological^1^** | 9,672 | 22.14% |
| **Breast** | 8,322 | 19.05% |
| **Genito-urinary^2^** | 7,624 | 17.46% |
| **Skin and melanoma** | 6,163 | 14.11% |
| **Gastrointestinal** | 4,124 | 9.44% |
| **Lung and thoracic^3^** | 2,104 | 4.82% |
| **Lower GastrointestinaI^4^** | 1,259 | 2.88% |
| **Other/unspecified** | 1,139 | 2.61% |
| **Gynaecological** | 939 | 2.15% |
| **Upper Gastrointestinal** | 632 | 1.45% |
| **Head and Neck** | 582 | 1.33% |
| **Endocrine** | 422 | 0.97% |
| **Central Nervous System** | 256 | 059% |
| **Sarcoma** | 158 | 0.36% |
| **Hepatobiliary** | 135 | 0.31% |
| **Pancreatic** | 118 | 0.27% |
| **Neuroendocrine** | 11 | 0.03% |
| **Bone/soft tissue** | 10 | 0.02% |
| **Cancer of Unknown Primary** | 6 | 0.01% |
|  | **43,676** |  |

**Supplementary Table S6.** Tumour prevalence across studies included.

^1^ acute myeloid leukaemia, chronic lymphocytic leukaemia, multiple myeloma, lymphoma, myelodysplastic syndrome, histiocytosis. ^2^ NSCLC, SCLC, thymoma, carcinoid, malignant pleural mesothelioma, thymic. ^3^ Kidney, bladder, adrenal, urothelial, testis, prostate, penile. ^4^ Colorectal and anal.
