## Supplementary material for "A comprehensive systematic review and meta-analysis of the global data involving 61,532 cancer patients with SARS-CoV-2 infection": Table S7

| **Publication** | **Total number of patients** | **Inpatient** | **Outpatient** | **Both (no breakdown)** | **Unknown/missing data** | **Possible nosocomial infection** | **Hospitalised at time of infection** |
| --- | --- | --- | --- | --- | --- | --- | --- |
| **Sharafeldin et al** | **38,614** | **19,515** | **19,099** |  |  |  |  |
| **Grivas et al** | **4966** | 2872 | 2094 |  |  | N |  |
| **Docherty et al** | **1743** | 1743 | - | - |  | N |  |
| **Özdemir et al** | **1523** | 862 | 661 |  |  |  |  |
| **Lievre et al** | **1289** | 734 | 555 |  |  | N |  |
| **Lee et al** | **1044** | 1044 | - | - |  | N |  |
| **Fratino et al** | **909** |  |  |  | 150 | N |  |
| **Pinato et al** | **890** | 760 | 130 | - |  | 155 |  |
| **Yigenoglu et al** | **740** |  |  | 740 |  |  |  |
| **Johannesen et al** | **547** | 104 | 443 |  |  |  |  |
| **Passamonti et al** | **536** | 536 | - | - |  | N |  |
| **Rüthrich et al** | **435** | 427 | 8 |  |  |  |  |
| **Montopoli et al** | **430** | - | - | 430 |  | N |  |
| **Robilotti et al** | **423** | 180 | 243 | - |  | N |  |
| **de Joode et al** | **351** | 351 | - | - |  | N |  |
| **Miyashita et al** | **334** | - | - | - | 334 | N |  |
| **Graeselli et al** | **331** | 331 |  | - |  | N |  |
| **Lunski et al** | **312** | 166 | 146 | - |  | N |  |
| **Jee et al** | **309** | 147 | 162 | - |  | N |  |
| **Song et al** | **248** | 248 | - | - |  | N |  |
| **COVIDSurg Collaborative** | **239** | 239 | - | - |  | N |  |
| **Tian et al** | **232** | 232 | - | - |  | N |  |
| **Di Cosimo et al** | **231** | 165 | 64 |  | 2 |  |  |
| **Mehta V et al** | **218** | 190 | 11 | - | 17 | 37/61 |  |
| **Yang K et al** | **205** | 205 | - | - |  | N |  |
| **Ferrari et al** | **198** | 88 | 110 |  |  |  |  |
| **Garassino et al** | **198** | 152 | 48 | - |  | 13/24 |  |
| **Mato et al** | **198** | 178 | 20 | - |  | N |  |
| **Scarfo et al** | **190** | 169 | 21 | - |  | N |  |
| **Mehta A et al** | **186** |  |  | 186 |  |  |  |
| **de Melo et al** | **181** | 181 | - | - |  | 83 |  |
| **Albiges et al** | **178** | 125 | 53 | - |  | 31 |  |
| **Martinez-Lopez et al** | **167** | 167 | - | - |  | N |  |
| **Russell et al** | **156** | 118 | 36 | - | 2 | N |  |
| **Basse et al** | **141** | 34 | 107 | - |  | N | 34 |
| **Lara et al** | **121** | 66 | 55 | - |  | N |  |
| **Brar et al** | **117** | 117 | - | - |  |  |  |
| **Angelis et al** | **113** | 101 | 12 | - |  | Not clear | 14 |
| **Gupta et al** | **112** | 112 | - | - |  | No |  |
| **Meng et al** | **109** | 109 | - | - |  | N |  |
| **Deng et al** | **107** | - | - | - | 107 | N |  |
| **Kabarriti et al** | **107** | 58 | 49 | - |  | N |  |
| **Zhang H et al** | **107** | 107 |  | - |  | N |  |
| **Dai et al** | **105** | 105 |  | - |  | N |  |
| **Luo et al** | **102** | 63 | 39 | - |  | N |  |
| **Cattaneo et al** | **102** | - |  | - | 102 | N |  |
| **Hultcrantz et al** | **100** | 75 | 25 | - |  | N |  |
| **Singh et al** | **85** | 73 | 12 | - |  | N |  |
| **Shah et al** | **80** | 80 | - | - |  |  |  |
| **Cook et al** | **75** | - | - | 75 |  | N |  |
| **Erdal et al** | **71** | 71 | - | - |  |  |  |
| **Sun et al** | **67** | 37 | 30 |  |  |  |  |
| **Booth et al** | **66** | 66 | - | - |  | N |  |
| **Yarza et al** | **63** | 63 | - | - |  | N |  |
| **Vuagnat et al** | **59** | 28 | 31 | - |  | 9 |  |
| **Wang B et al** | **58** | 36 | 22 | - |  | N |  |
| **Assaad et al** | **55** | - |  | - | 55 | N |  |
| **Fox et al** | **55** | 47 | 8 | - |  | 4 |  |
| **Yang F et al** | **52** | - |  | - | 52 | N |  |
| **Suleyman et al** | **49** | 43 | 6 | - | - | N |  |
| **Rogado et al** | **45** | 38 | 7 | - | - | P |  |
| **Sanchez-Pina et al** | **39** | - | - | 39 | - | 1 | 1 |
| **Ma et al** | **37** | 37 | - | - | - | N |  |
| **Aries et al** | **35** | 35 | - | - | - | N |  |
| **Martin-Moro et al** | **34** | 34 | - | - | - | N |  |
| **Joharatnam-Hogan et al** | **30** | 30 | - | - | - | 1 |  |
| **Zhang L et al** | **28** | 28 | - | - | - | 8 |  |
| **Kalinsky et al** | **27** | 7 | 20 | - | - | N |  |
| **Malard et al** | **25** |  |  | 25 | - | 10 possibly | 6 |
| **Stroppa et al** | **25** | 25 |  |  | - | N |  |
| **Ciceri et al** | **22** | 22 |  |  | - | N |  |
| **Tomlins et al** | **20** | 20 |  |  | - | N |  |
| **Bogani et al** | **19** | 19 |  |  | - | N |  |
| **Liang et al** | **18** | 18 |  |  | - | N |  |
| **Guan et al** | **18** | 18 |  |  | - | P |  |
| **Tagliamento et al** | **17** | 13 | 4 |  | - | N |  |
| **Wang L et al** | **15** | 15 |  |  | - | N |  |
| **He et al** | **13** | 13 |  |  | - | N |  |
| **Lattenist et al** | **13** | 13 |  |  | - | 1 |  |
| **Yu et al** | **12** | 12 |  |  | - | N |  |
| **Wu et al** | **11** |  |  |  | 11 | N |  |

**Supplementary Table S7.** Inpatient vs. outpatient care across 73 studies as well as possible or probable nosocomial infection.
