## Supplementary material for "A comprehensive systematic review and meta-analysis of the global data involving 61,532 cancer patients with SARS-CoV-2 infection": Table S8

| **Publication** | **Imaging Modality** | **Bilateral involvement** | **Ground glass** | **Patchy shadows** | **Consolidation** | **Infiltrates** | **Other*** |
| --- | --- | --- | --- | --- | --- | --- | --- |
| **Pinato et al** | CT or CXR: 811 | 427 |  |  |  |  |  |
| **Robilotti et al** | CT: 39, CXR: 207 | 23 | 30 |  | 13 |  | 32 |
| **Jee et al** |  | 76 |  |  | 34 |  |  |
| **Song et al** |  | 188 |  |  | 53 |  | 185 |
| **Tian et al** | CT: 232 | 150 | 148 | 126 |  |  | 165 |
| **Yang K et al** | CT: 190 | 173 | 132 |  |  |  |  |
| **Mato et al** | CT or CXR: 183 |  |  |  |  |  |  |
| **Albiges et al** | CT: 133 |  |  |  |  |  |  |
| **Basse et al** | CT: 80 |  | 24 |  | 7 |  |  |
| **Angelis et al** | CT: 20, CXR: 86 |  |  |  |  | 47 |  |
| **Meng et al** |  | 27 | 25 |  |  |  |  |
| **Cook et al** |  |  |  |  |  | 56 |  |
| **Yarza et al** |  | 33 |  |  |  | 60 |  |
| **Vuagnat et al** | CT: 39 |  | 14 |  |  |  |  |
| **Fox et al** | CT or CXR: 9 |  | 71 |  |  |  | 7 |
| **Sanchez-Pina et al** | CT: 39 | 28 |  |  |  |  |  |
| **Martin-Moro et al** |  |  |  |  |  | 30 |  |
| **Joharatnam-Hogan et al** | CXR: 29 |  |  |  |  |  |  |
| **Zhang L et al** | CT: 28 |  | 21 |  | 13 |  | 4 |
| **Kalinsky et al** | CT: 39 |  |  |  |  |  |  |
| **Malard et al** | CT: 14, CXR: 7 | 21 |  |  |  |  |  |
| **Stroppa et al** | CT: 22, CXR: 3 | 22 |  |  |  |  | 25 |
| **Bogani et al** | CT: 18 |  |  |  |  |  |  |
| **Guan et al** | CT: 14, CXR: 7 |  |  |  |  |  |  |
| **He et al** | CT: 22, CXR: 3 |  | 8 | 2 | 1 |  | 1 |
| **Lattenist et al** | CT: 18 |  |  |  |  |  |  |
| **Yu et al** | CT: 11 |  |  |  |  |  |  |
| **Total:** | CT: 958, CXR: 525, either: 1,003 | 1,168 | 473 | 128 | 121 | 193 | 419 |

**Supplementary Table S8.** Chest radiograph ± chest x ray imaging on admission with radiological changes documented, where reported**.** *Nodules/interstitial thickening/erratic paving
