## Supplementary material for "A comprehensive systematic review and meta-analysis of the global data involving 61,532 cancer patients with SARS-CoV-2 infection": Table S9

| **A) Age (obs: 14)** |  |  |  |  | **tau2=0.06352, I^2^=54.7%** |
| --- | --- | --- | --- | --- | --- |
| *Effect Size* | *exp (b)* | *Standard Error* | *t* | *P >\|t\|* | *[95% Confidence Interval]* |
| Age | 0.9572513 | 0.0166774 | -2.51 | 0.028 | 0.9215953 – 0.9942868 |
| **B) Male cancer patients (obs: 17)** |  |  |  |  | **tau2=0.08595, I^2^=67.92%** |
| *Effect Size* | *exp (b)* | *Standard Error* | *t* | *P >\|t\|* | *[95% Confidence Interval]* |
| Male cancer patients | 1.187946 | 0.935615 | 0.22 | 0.830 | 0.221688 – 6.365771 |
| **C) Age and male cancer patients (obs: 14)** |  |  |  |  | **tau2=0.06999, I^2^=57.07%** |
| *Effect Size* | *exp (b)* | *Standard Error* | *t* | *P >\|t\|* | *[95% Confidence Interval]* |
| Age | 0.9517435 | 0.0188232 | -2.50 | 0.029 | 0.9112026 – 0.994088 |
| Male cancer patients | 3.526408 | 7.123145 | 0.62 | 0.545 | 0.0413537 – 300.7112 |

**Supplementary Table S9** Meta-regression results on the impact of (A) age, (B) male gender, and (C) both age and male gender.

Obs, observations.
