## Supplementary material for "A comprehensive systematic review and meta-analysis of the global data involving 61,532 cancer patients with SARS-CoV-2 infection": Table S10

|  | Cancer covid patients | Not hospitalised/  unknown | Number hospitalised | ICU admission | IMV | CRRT | ECMO | Haemo-  dialysis | IV  Vasopressors | Transition to  DNR/DNI | Discharged | Remain  Hospitalised | Death |
| --- | --- | --- | --- | --- | --- | --- | --- | --- | --- | --- | --- | --- | --- |
| **Sharafeldin et al, 2021** | **38,614** | 19,099 | 19,515 |  | 1,600 |  | 27 |  |  |  |  |  | 2,888 |
| **Grivas et al, 2020** | **4,966** | 2,094 | 2,872 | 232 | 292 |  |  |  |  |  |  |  | 695 |
| **Özdemir et al, 2020** | **1,523** | 661 | 862 | 173 | 113 |  |  |  |  |  |  |  | 77 |
| **Lievre et al, 2020** | **1289** | 555 | 734 | 110 | 49 |  |  |  |  |  |  |  | 322 |
| **Lee et al, 2020** | **1044** | 1003 | 41 | 63 | 43 |  |  |  |  |  |  | 41 | 295 |
| **Pinato et al, 2020** | **890** | 120 | 760 | 110 | 35 |  |  |  |  |  |  |  | 299 |
| **Yigenoglu et al, 2020** | **740** | 452 | 288 | 140 | 102 |  |  |  |  |  |  |  | 102 |
| **Johannesen et al, 2020** | **547** | 427 | 120 | 17 | 17 |  |  |  |  |  |  |  | 56 |
| **Passamonti et al, 2020** | **536** | 85 | 451 | 82 |  |  |  |  |  |  | 242 | 11 | 198 |
| **Rüthrich et al, 2020** | **435** | 8 | 427 | 119 | 78 |  |  |  |  |  | 292 | 46 | 97 |
| **Robilotti et al, 2020** | **423** | 243 | 180 | 48 | 40 |  |  |  |  |  |  |  | 51 |
| **Miyahsita et al, 2020** | **334** |  |  |  | 37 |  |  |  |  |  |  |  | 37 |
| **Lunski et al, 2020** | **312** | 146 | 166 | 48 |  |  |  |  |  |  |  |  | 66 |
| **Jee et al, 2020** | **309** | 162 | 147 | 43 |  |  |  |  |  |  |  |  | 31 |
| **Tian et al, 2020** | **232** | 0 | 232 |  | 21 |  |  |  |  |  |  |  | 46 |
| **Di Cosimo et al, 2020** | **231** | 66 | 165 | 12 | 19 |  |  |  |  |  |  |  | 81 |
| **Mehta V et al, 2020** | **218** | 28 | 190 | 23 | 45 |  |  |  |  |  |  | 35 | 61 |
| **Yang K et al, 2020** | **205** | 0 | 205 | 30 | 21 | 5 |  | 5 |  |  | 165 |  | 40 |
| **Ferrari et al, 2020** | **198** | 110 | 88 | 37 |  |  |  |  |  |  | 149 | 16 | 33 |
| **Garassino et al, 2020** | **198** | 48 | 152 | 13 | 9 |  |  |  |  |  |  | 92 | 66 |
| **Mato et al, 2020** | **198** | 20 | 178 | 68 | 53 |  |  |  | 47 |  | 63 | 49 | 66 |
| **Scarfo et al, 2020** | **190** | 21 | 169 | 39 |  |  |  |  |  |  | 96 | 37 | 56 |
| **Mehta A et al, 2020** | **186** |  |  |  | 12 |  |  |  |  |  |  |  | 27 |
| **de Melo et al, 2020** | **178** | 53 | 125 | 80 |  |  |  |  |  |  |  |  | 60 |
| **Albiges et al, 2020** | **167** | 0 | 167 |  | 15 |  | 16 |  |  |  | 80 | 16 | 31 |
| **Martinez-Lopez et al, 2020** | **167** | 0 | 167 |  | 15 |  |  |  |  |  | 110 | 1 | 56 |
| **Russell et al, 2020** | **156** | 38 | 118 | 13 |  |  |  |  |  |  |  |  | 34 |
| **Basse et al, 2020** | **141** | 91 | 50 | 11 |  |  |  |  |  |  | 100 | 11 | 26 |
| **Lara et al, 2020** | **121** | 55 | 66 | 20 | 9 |  |  |  |  |  | 39 | 11 | 18 |
| **Montopoli et al, 2020** | **118** | 8 | 78 | 14 |  |  |  |  |  |  |  |  | 18 |
| **Brar et al, 2020** | **117** | 117 | 0 |  | 4 |  |  |  |  |  | 85 | 3 | 29 |
| **Angelis et al, 2020** | **113** | 12 | 101 | 12 | 8 |  |  |  |  |  |  |  | 29 |
| **Meng et al, 2020** | **109** | 0 | 109 |  | 9 |  |  |  |  |  | 77 |  | 32 |
| **Zhang H et al, 2020** | **107** | 0 | 107 |  | 18 |  |  |  |  |  | 84 |  | 23 |
| **Dai et al, 2020** | **105** | 0 | 105 | 20 | 11 | 4 | 3 | 4 |  |  |  |  | 12 |
| **Luo et al, 2020** | **102** | 39 | 63 | 21 | 18 |  |  |  |  | 18 | 53 | 24 | 25 |
| **Cattaneo et al, 2020** | **102** |  |  | 21 | 5 |  |  |  |  |  |  |  | 40 |
| **Hultcrantz et al, 2020** | **100** | 25 | 75 | 17 | 13 |  |  |  |  |  |  |  | 22 |
| **Singh et al, 2020** | **85** | 12 | 73 | 30 | 23 | 7 | 30 |  | 19 | 39 | 38 | 3 | 32 |
| **Shah et al, 2020** | **80** | 0 | 80 |  |  |  |  |  |  |  |  |  |  |
| **Cook et al, 2020** | **75** | 3 | 72 | 9 | 6 |  |  |  |  |  |  |  | 41 |
| **Erdal et al, 2020** | **71** | 0 | 71 | 18 | 17 |  |  |  |  |  |  |  |  |
| **Sun et al, 2020** | **67** | 30 | 37 | 17 |  |  |  |  |  |  |  |  | 9 |
| **Booth et al, 2020** | **66** | 0 | 66 |  | 3 |  |  |  |  |  | 28 | 4 | 34 |
| **Yarza et al, 2020** | **63** | 0 | 63 | 0 | 0 |  |  |  |  |  |  |  | 16 |
| **Vuagnat et al, 2020** | **59** | 31 | 28 | 4 | 1 |  |  |  |  |  | 45 | 10 | 4 |
| **Wang B et al, 2020** | **58** | 22 | 36 | 7 | 5 |  |  |  |  |  | 22 | 1 | 14 |
| **Fox et al, 2020** | **55** | 4 | 51 |  | 25 | 1 |  |  | 5 |  | 35 | 1 | 19 |
| **Yang F et al, 2020** | **52** | 0 | 52 |  | 0 | 1 |  |  |  |  | 41 |  | 11 |
| **Suleyman et al, 2020** | **49** | 6 | 43 | 23 |  |  |  |  |  |  |  |  | 19 |
| **Rogado et al, 2020** | **45** | 7 | 38 |  |  |  |  |  |  |  |  | 26 | 19 |
| **Sanchez-Pina et al, 2020** | **39** | 5 | 34 | 1 |  |  |  |  |  |  |  |  | 14 |
| **Aries et al, 2020** | **35** |  |  |  |  |  |  |  |  |  | 21 | 1 | 14 |
| **Martin-Moro et al, 2020** | **34** | 0 | 34 | 2 | 4 |  |  |  |  |  |  | 5 | 11 |
| **Joharatnam-Hogan et al, 2020** | **30** | 0 | 30 |  |  |  |  |  |  |  |  |  | 11 |
| **Zhang L et al, 2020** | **28** | 0 | 28 | 6 | 12 |  |  |  |  |  | 10 |  | 8 |
| **Kalinsky et al, 2020** | **27** | 20 | 7 |  |  |  |  |  |  |  | 26 | 0 | 1 |
| **Malard et al, 2020** | **25** | 0 | 25 |  | 6 |  |  |  |  |  |  |  | 9 |
| **Stroppa et al, 2020** | **25** | 0 | 25 | 1 |  |  |  |  |  |  |  |  | 9 |
| **Ciceri et al, 2020** | **22** | 0 | 22 |  |  |  |  |  |  |  | 11 | 0 | 11 |
| **Bogani et al, 2020** | **19** | 0 | 19 | 2 |  |  |  |  |  |  | 14 | 2 | 3 |
| **Liang et al, 2020** | **18** | 0 | 18 | 7 |  |  |  |  |  |  |  |  | 7 |
| **Guan et al, 2020** | **18** | 0 | 18 | 5 | 2 |  |  |  |  |  |  |  | 3 |
| **Tagliamento et al, 2020** | **17** | 4 | 13 |  |  |  |  |  |  |  |  | 13 | 4 |
| **He et al, 2020** | **13** | 0 | 13 |  | 1 |  |  |  |  |  |  | 0 | 8 |
| **Lattenist et al, 2020** | **13** | 0 | 13 | 2 | 1 |  |  |  |  |  |  | 0 | 6 |
| **Yu et al, 2020** | **12** | 0 | 12 |  |  |  |  |  |  |  | 9 | 3 | 3 |
| **Wu et al, 2020** | **11** | 4 | 7 |  |  |  |  |  |  |  |  | 1 | 4 |

**Supplementary Table S10** Patient outcomes in 68 studies.

CRRT, continuous renal-replacement therapy; DNR/DNI, do not resuscitate/do not intubate; ECMO, extracorporeal membrane oxygenation; ICU, intensive care unit; IMV, invasive mechanical ventilation
