## Supplementary material for "A comprehensive systematic review and meta-analysis of the global data involving 61,532 cancer patients with SARS-CoV-2 infection": Table S11

| **Publication** | **Duration of hospital stay (days)** |
| --- | --- |
| Pinato et al | 10 (IQR 5-18) |
| Passamonti et al | 16 (IQR 9-29) |
| Lunski et al | 6 (IQR 3-15) |
| Song et al | 22 |
| Yang K et al | 19 (IQR 12-33) |
| Garassino et al | 12 |
| Albiges et al | 10 (IQR 1-40) |
| Lara et al | 7 (IQR 4-10) |
| Dai et al* | 27.01 (SD 9.52) |
| Singh et al | 9.5 (IQR 1-27) |
| Erdal et al | 10 (1-39) |
| Yarza et al* | 9.2 (IQR 8.17-10.33) |
| Wang B et al | 22 |
| Fox et al | 13 (IQR 0-135) |
| Zhang L et al | 19 (IQR 16-28.5 |

**Supplementary Table S11.** Median duration of hospital stay (days).

*Mean duration.
