## Supplementary material for "A comprehensive systematic review and meta-analysis of the global data involving 61,532 cancer patients with SARS-CoV-2 infection": Table S12

| **Publication** | **Mortality in those with cancer and COVID-19** | **Cancer COVID patients** | **%** | **Median follow up, days (range) Median Mean** | |
| --- | --- | --- | --- | --- | --- |
| Sharafeldin et al | 3,164 | 38,614 | 8.19% | - | |
| Grivas et al | 695 | 4966 | 14.0% | 42 (22-90) |  |
| Docherty et al | 617 | 1743 | 35.4% | - | |
| Özdemir et al | 77 | 1523 | 5.06% | 50 (1-74) |  |
| Lièvre et al | 322 | 1289 | 24.98% | 34 (32-36) |  |
| Lee et al | 295 | 1044 | 28.3% | 6 (2-11) |  |
| Fratino et al | 150 | 909 | 16.5% | - |  |
| Pinato et al | 299 | 890 | 33.6% |  | 19 (+/-16.3) |
| Yigenoglu et al | 102 | 740 | 13.78% |  |  |
| Johannesen et al | 56 | 547 | 10.24% |  |  |
| Passamonti et al | 198 | 536 | 36.9% | 20 (10-34) |  |
| Rüthrich et al | 97 | 435 | 22.30% |  |  |
| Montopoli et al | 75 | 430 | 17.4% | - |  |
| Robilotti et al | 51 | 423 | 12.1% | - |  |
| de Joode et al | 114 | 351 | 32.5% | - |  |
| Miyashita et al | 37 | 334 | 11.1% | - |  |
| Graeselli et al | 202 | 331 | 61.0% | 69 (60-78) |  |
| Lunski et al | 66 | 312 | 21.1% | - |  |
| Jee et al | 31 | 309 | 10.0% | - |  |
| Song et al | 40 | 248 | 16.1% | - |  |
| COVIDSurg Collaborative | 66 | 239 | 27.6% | - |  |
| Tian et al | 46 | 232 | 19.8% | 29 (22-38) |  |
| Di Cosimo et al | 81 | 231 | 35.06% | 138 (12-218) |  |
| Mehta V et al | 61 | 218 | 28.0% | - |  |
| Yang K et al | 30 | 205 | 14.6% | 68 (59-78) |  |
| Ferrari et al | 33 | 198 | 16.67% | 61 |  |
| Garassino et al | 66 | 198 | 33.3% | 15 (8-24) |  |
| Mato et al | 66 | 198 | 33.3% | 16 (1-43) |  |
| Scarfo et al | 56 | 190 | 29.5% | 23 (2-86) |  |
| Mehta A et al | 27 | 186 | 14.52% | 63 |  |
| de Melo et al | 69 | 181 | 38.1% | 5 (2-10.3) |  |
| Albiges et al | 31 | 178 | 17.4% | 23 (13-33) |  |
| Martinez-Lopez et al | 56 | 167 | 33.5% | - |  |
| Russell et al | 34 | 156 | 21.8% | 37 (18-49) |  |
| Basse et al | 26 | 141 | 18.4% | - |  |
| Lara et al | 17 | 121 | 14.0% | - |  |
| Brar et al | 29 | 117 | 24.8% |  |  |
| Angelis et al | 29 | 113 | 25.7% | - |  |
| Gupta et al | 60 | 112 | 53.6% | 16 (8-28) |  |
| Meng et al | 32 | 109 | 29.4% | - |  |
| Deng et al | 6 | 107 | 5.6% | - |  |
| Kabarriti et al | 24 | 107 | 22.4% | 7 ( 0.5-39) |  |
| Zhang H et al | 23 | 107 | 21.5% | - |  |
| Dai et al | 12 | 105 | 11.4% | - |  |
| Luo et al | 25 | 102 | 24.5% | 25 (10-36) |  |
| Cattaneo et al | 40 | 102 | 39.2% | - |  |
| Hultcrantz et al | 22 | 100 | 22.0% | - |  |
| Singh et al* | 32 | 85 | 37.6% | 31 (20-36) |  |
| Shah et al | 31 | 80 | 38.8% | - |  |
| Cook et al | 41 | 75 | 54.7% | - |  |
| Erdal et al | 17 | 71 | 23.9% | - |  |
| Sun et al | 9 | 67 | 13.4% | - |  |
| Booth et al | 34 | 66 | 51.5% | 32.5 |  |
| Yarza et al | 16 | 63 | 25.4% | - |  |
| Vuagnat et al | 4 | 59 | 6.8% | - |  |
| Wang B et al | 14 | 58 | 24.1% | - |  |
| Assaad et al | 8 | 55 | 14.5% | 25 |  |
| Fox et al | 19 | 55 | 35.0% | 27 (17-43) |  |
| Yang F et al | 11 | 52 | 21.2% | - |  |
| Suleyman et al | 19 | 49 | 38.8% | - |  |
| Rogado et al | 19 | 45 | 42.2% | 14 (1-28) |  |
| Sanchez-Pina et al | 14 | 39 | 35.9% | - |  |
| Ma et al | 5 | 37 | 13.5% | - |  |
| Aries et al | 14 | 35 | 40.0% | - |  |
| Martin-Moro et al | 11 | 34 | 32.4% | 26 |  |
| Joharatnam-Hogan et al | 6 | 30 | 20.0% | - |  |
| Zhang L et al | 8 | 28 | 28.6% | - |  |
| Kalinsky et al | 1 | 27 | 3.7% | 26 (1-38) |  |
| Malard et al | 9 | 25 | 36.0% | 29 (14-40) |  |
| Stroppa et al | 9 | 25 | 36.0% | - |  |
| Ciceri et al | 11 | 22 | 50.0% | 14 (7-25) |  |
| Tomlins et al | 3 | 20 | 15.0% | - |  |
| Bogani et al | 3 | 19 | 15.8% | - |  |
| Liang et al | 7 | 18 | 38.9% | - |  |
| Guan et al | 3 | 18 | 16.7% | 10 (8-14) |  |
| Tagliamento et al | 4 | 17 | 23.5% | 15 |  |
| Wang L et al | 3 | 15 | 20.0% | - |  |
| He et al | 8 | 13 | 61.5% | - |  |
| Lattenist et al | 6 | 13 | 46.2% | - |  |
| Yu et al | 3 | 12 | 25.0% | - |  |
| Wu et al | 4 | 11 | 36.4% | - |  |

**Supplementary Table S12** Mortality of cancer patients reported across studies.

* Death defined mortality + transfer to hospice, not reported separately.
