## Supplementary material for "A comprehensive systematic review and meta-analysis of the global data involving 61,532 cancer patients with SARS-CoV-2 infection": Table S13

| **Type of cancer treatment** | **Number of studies included in analysis** | **Number of patients included across studies (range)** | **Pooled case fatality rate** |
| --- | --- | --- | --- |
| Surgery | 7 | 4 to 56 | 19% |
| Chemotherapy | 22 | 3 to 802 | 31% |
| Endocrine | 9 | 4 to 483 | 11% |
| Immunotherapy | 14 | 3 to 248 | 22% |
| Radiotherapy | 9 | 2 to 95 | 20% |
| Targeted therapy | 15 | 1 to 693 | 18% |

**Supplementary Table S13** Pooled case fatality rates for various cancer treatments.
