## Supplementary material for "A comprehensive systematic review and meta-analysis of the global data involving 61,532 cancer patients with SARS-CoV-2 infection": Table S14

| **Name of Study** | **Trial or Registration Number** | **Location** | **Number of patients and patient characteristics** | **Primary endpoint** |
| --- | --- | --- | --- | --- |
| CovidSurg – Cancer | NCT04384926 | Global | 1000 patients planned for cancer surgery | 30-day postoperative COVID-19 infection rate.  https://globalsurg.org/cancercovidsurg/ |
| COVID-19 and Cancer Consortium Registry (CCC19) | NCT04354701 | Global | 10,000 patients | Collect data about cancer patients who have been infected with COVID-19 via web-based REDCap Survey.  https://ccc19.org/ |
| American Society of Haematology Research Collaborative (ASH RC) COVID-19 Registry for Hematologic Malignancy | - | Global | - | Captures data on patients with COVID-19 and have been or are currently being treated for hematologic malignancy.  https://www.ashresearchcollaborative.org/covid-19-registry |
| ASCO Survey on COVID-19 in Oncology (ASCO) Registry | NCT04659135 | USA | 2000 patients | Changes to Cancer Treatments. |
| Thoracic cancers international COVID-19 collaboration (TERAVOLT) | - | Global | - | A global consortium designed to gather information on patients with thoracic cancer infected with COVID-19 regardless of therapies administered.  http://www.etop-eu.org/index.php?option=com_content&view=article&id=115644&catid=13&Itemid=557 |
| Clinical Characterisation Protocol-Cancer-UK | NCT04603105 | UK | 9000 patients | To determine the COVID-19 fatality rate overall in the cancer population using the most up to date dataset from the first wave as well as to determine the COVID-19 fatality rate in different tumour types. https://isaric.tghn.org/UK-CCP. |
| UK Coronavirus Cancer Monitoring Project |  | UK |  | The UK Coronavirus cancer monitoring scheme is a clinician-led reporting project recoding data related to cancer patients who have tested positive for COVID-19 across the UK.  <https://ukcoronaviruscancermonitoring.com/> |
| ONCOVID: | NCT04393974 | UK | 1000 patients | Describe presenting characteristics and severity or SARS-CoV-2 infection in patients with cancer. To assess what factors are involved in prognosis of cancer patients with COVID-19.  https://www.oncovid.net/ |
| Prospective Analysis of Morbi-mortality of Patients With Cancers in Active Phase of Treatment Suspected or Diagnosed of a SARS-CoV-2 Infection (ONCOVID-19) | NCT04363632 | France | 1231 patients | Mortality of cancer patients under active anticancer treatment. |
| UK Covid and gynaecological cancer study (UKCOGS-UK) |  | UK |  | Evaluate the MDT decision making for gynaecological cancer, patient outcomes across the UK in response to the COVID-19 pandemic.  |
| PACE: Patients with AML and COVID-19 Epidemiology | 282870 (IRAS ID) | UK | 100 patients with AML | Record how many patients have had COVID-19 previously, have an active infection or go on to develop COVID-19 whilst receiving treatment for their AML. |
| Covid-19 in Hematological Malignancies (EPICOVIDEHA) | NCT04733729 | Italy | 3000 patients with haematological malignancies | Epidemiology of COVID-19 infection in patients with haematological malignancies |
| Screening and Identification of SARS-CoV-2 Infection and Progression in Cancer Patients Based on Bioinformatics Analysis | ChiCTR2000030807 | China | 100 patients. | Clinical characteristics and prognosis of cancer patients with novel coronavirus pneumonia (COVID-19) based on bioinformatics analysis.  http://www.chictr.org.cn/showproj.aspx?proj=51019 |
| COVID-19 Infection and Multiple Myeloma (EMN-COVID) | NCT04492371 | Global | 500 patients with multiple myeloma and COVID-19 infection | Nature of COVID19, costs related to COVID-19, systemic anti-cancer therapy subgroup, laboratory values collected at hospitalization, COVID-19 infection in myeloma patient subgroups, incidence of COVID-19 infection in frail patients and infection outcome in different countries. |
| LunG and Melanoma canceR pAtients coVId19 Disease (GRAVID) | NCT04344002 | Spain | 200 patients with lung cancer and COVID-19 | Clinical data of lung cancer patients with COVID-19 diagnoses, diagnosis data, treatments received and prognostic factors. |
| COVID-19 Infection in Cancer Patients (COICA) | NCT04569292 | Italy | 150 patients | Describe cancer patients with COVID-19 and their clinical course |
| Registry on NEN Patients and COVID-19 | NCT04444401 | Italy | 50 patients with neuroendocrine tumours | Correlation between clinical parameters and SARS-CoV-2 infection as well as clinical outcome of SARS-CoV-2 Infection |
| Effects of COVID-19 Pandemic on the Diagnosis and Outcomes of Colorectal Cancer (COVID-CRC) (COVID-CRC) | NCT04712292 | Italy | 2000 patients | Oncologic stage according to TNM classification. |
| NCI COVID-19 in Cancer Patients, NCCAPS Study | NCT04387656 | Global | 2000 patients | Patient variables (factors) associated with severe acute respiratory syndrome (SARS) coronavirus 2 (COVID-19) severity, effects of COVID-19 on cancer therapy and association with clinical outcomes and physical health (patient-reported health-related quality of life). |
| Prospective Determination of COVID-19 Infection Rate in a Chemotherapy Unit in Mexico | NCT04567979 | Mexico | 149 patients with solid malignant disease and healthcare workers | SARS-CoV-2 infection rate in patients with solid tumours. |
| A Study of Risk Factors for the COVID-19 Virus Infection | NCT04697927 | USA | 10,000 patients | To develop a comprehensive registry database. |
| Registry of Patients With Hematologic Disease and COVID-19 in Russia (CHRONOS19) | NCT04422470 | Russia | 200 patients with malignant and non-malignant haematological disease | 30-day all-cause mortality |
| Long-term Evolution of Pulmonary Involvement of Novel SARS-COV-2 Infection (COVID-19): Follow the Covid Study | NCT04605757 | Italy | 100 patients | Long term evolution of clinical involvement due to SARS-COV-2 pneumonia / symptoms, respiratory rate, blood gas exchange parameters, pulmonary function tests,  ogical involvement. |
| Outcome of cancer patients infected with COVID-19, including toxicity of cancer treatments | - | France | 7,251 patients | Clinical deterioration, defined as the need for O2 supplementation of 6l/min or more, or death of any cause. |
| Incidence of thrombosis and hemorrhage in hospitalized cancer patients with COVID-19 |  | USA | 45 cancer patients and 353 non cancer patients | Evaluate cumulative incidences of thrombotic and hemorrhagic events in hospitalized COVID-19 patients with and without active cancer at 28 days. |
| Poor outcome and prolonged persistence of SARS-CoV-2 RNA in COVID-19 patients with  haematological malignancies; King’s College Hospital experience | - | UK | 80 patients | Comparison of first 80 patients with haematological malignancy with all other  patients admitted to our hospital with COVID-19 in the same time frame to define  relative risk and identify factors that increase mortality within this subgroup. |
| Nosocomial outbreak of SARS-CoV-2 infection in a haematological unit – High mortality rate in infected patients with haematologic malignancies | - | Poland | 19 patients with haematological malignancies and 20 health care workers | Compare morbidity and mortality in infected and non-infected patients after exposure to SARS-CoV-2. |
| COVID-19 infection in hematopoietic cell transplantation: age, time from transplant and steroids matter | - | USA | 34 haematopoietic stem cell transplant recipients | To identify mortality risk to HCT recipients |
| COVID‐19 in patients with hematological malignancies: A retrospective case series | - | Spain | 41 patients with haematological malignancies | Characterize the real impact of COVID‐19 in patients with hematological neoplasms, in order to optimize clinical decision‐making. |
| Clinical characteristics and outcome of multiple myeloma patients with concomitant COVID-19 at Comprehensive Cancer Centers in Germany | - | Germany | 21 patients | Characterize a population of MM patients registered from 10 institutions who developed COVID-19 at hotspot areas in Germany |
| COVID-19, impact on myeloma patients | - | Belgium | 20 patients with multiple myeloma | Assess the impact of COVID-19 in the Belgian MM community, |
| **Psychological Impact:** |  |  |  |  |
| COVID-19 Pandemic Impact on Patients With Cancer - a Danish Survey (COPICADS) | NCT04389996 | Denmark | 5000 patients | Overall Quality of Life |
| Impact of the COVID-19 Pandemic and HRQOL in Cancer Patients and Survivors | NCT04447222 | USA | 1242 patients | Coronavirus disease-2019 (COVID19)-specific psychological distress |
| Impact of COVID-19 on Lung Cancer Patients | NCT04538456 | UK | 800 patients with lung cancer | Physical, social impact and psychological impact |
| Perception of the COVID-19 Pandemic in Patients With Haematological or Solid Neoplasias | NCT04649320 | Austria | 300 patients with malignant disease (solid or haematological) | Influence of the COVID-19 pandemic on cancer patient's daily life |
| The effects of prevention and control measures on treatment and psychological status of cancer patients during the novel coronavirus pneumonia (COVID-19) outbreak | ChiCTR2000030686 | China | 300 patients with malignant disease | The effects of prevention and control measures on treatment and psychological status of cancer patients during the novel coronavirus pneumonia (COVID-19) outbreak  http://www.chictr.org.cn/showproj.aspx?proj=50714 |

**Supplementary Table S14** Table of ongoing and further cancer observational studies related to the SARS-CoV-2/COVID-19 pandemic in patients with malignant disease.

AML, acute myeloid leukaemia.
